# Modeling COVID-19 in different countries as sequences of SI waves

**DOI:** 10.1101/2022.11.22.22282631

**Authors:** Rainer Janssen, Juergen Mimkes

## Abstract

The COVID-19 pandemic has been a huge challenge worldwide for many institutions, researchers, national health organizations, and the pharmaceutical industry. As natural scientists and engineers, we attempted to contribute by calculating models and analyzing data to keep track of the pandemic.

While a frequent goal is to predict the next pandemic wave by considering all influencing parameters, we examined methods to calculate a model course of the entire pandemic. This is done by reconstructing the course of infections into multiple model waves that sum up into a pandemic model that is close to the real course. The model wave parameters are varied by an algorithm, such as the Excel solver, to minimize the difference between the real and model courses.

By reconstructing the course of infections using the commonly known SIR model, we found that the calculated model parameters were ambiguous and difficult to interpret. In contrast, we found that sequenced SI model waves provide an astonishing precise digital representation of the pandemic course. Until November 2022, we found between six and 16 waves (depending on the country) in each of the 14 countries investigated.

The calculated parameters are easy to interpret and are comparable between different waves and countries. These wave parameters may be correlated with the virus types and measures in each country by other researchers. New waves are detectable early as they show a certain deviation from the actual model wave. After the maximum of the last real wave, the model indicates the further procedure for the pandemic course.

## Introduction

Modeling epidemic courses began with differential equations and probability calculations by Ross [1] and Kermack and McKendrick [2]. They developed the SI model of infections that considers only infected (I) and susceptible (S) persons, and later, the SIR model, which also includes recovered (R) persons.

Johns Hopkins University (JHU, [3]) has published data on global COVID-19 infections since January 2020. When the first data of the new epidemic in China were available on the Internet, we started to model John’s Hopkins data according to the SI model of epidemic growth.

Later on, we have started to use data by the organization Our World in Data (OWID, [4]) as international input data. For Germany, we use data by the Robert-Koch-Institute (RKI, [5]).

We varied the SI model parameters to minimize the differences between the real and model courses. In the first wave, this was performed manually; later, the high number of parameters led us to use the Excel solver tool with the GRG nonlinear method.

### The SI model

We analyzed the data using the simple SI infection model of a population N that contains susceptible S(t) and infected people I(t):

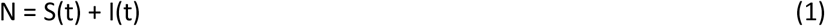

The number of daily newly infected people n(t) at time t depends on the infection rate (b),

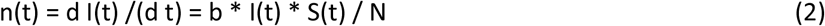

Equation (2) may be solved by a closed mathematical term, the sum of all infected people I(t) at time t is

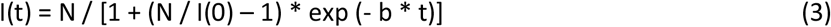

I(0) is the initial number of infected people, t = 0. Solution (3) contains two unknown parameters: the total number of susceptible people, N, and infection rate, b. These constants were easily obtained from the logarithmic plot of the infection data, as shown in Fig. 1.

**Fig. 1.**
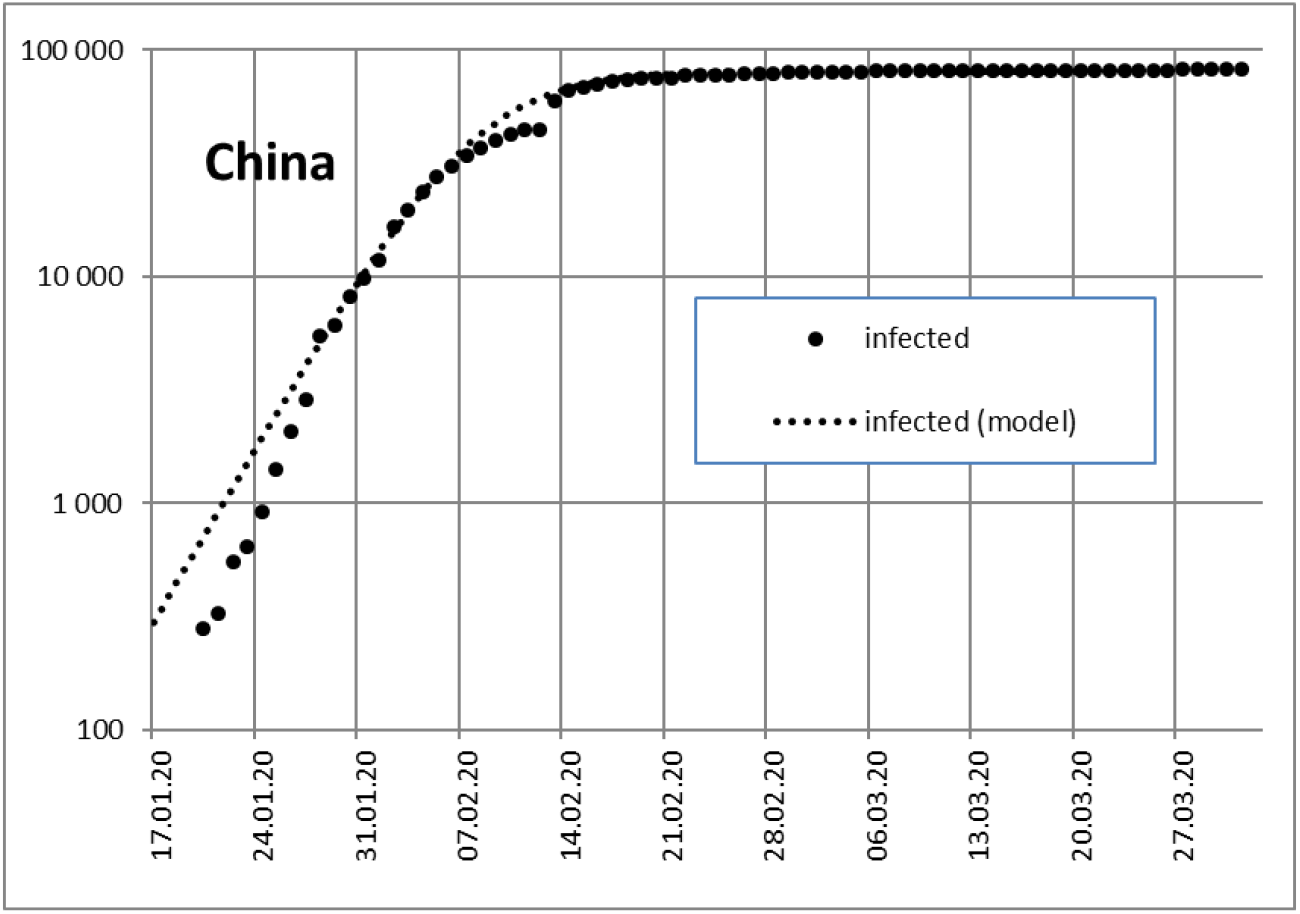
COVID-19 infection I(t) in China and SI model (logarithmic) [6]

China was the first country to be hit by COVID-19. Initially, the fit of the SI model was in good agreement with COVID-19 data in China, as shown in Fig. 1. The two unknown constants b and N were obtained from the slope and maximum in the logarithmic plot in Fig. 1.

In Europe, Italy was the first country to be hit by COVID-19. When we modeled the course of infections in Italy after April 2020, the simple SI model in Fig. 2 did not fit the real course because the SI model only allowed for symmetrical model waves.

**Fig. 2.**
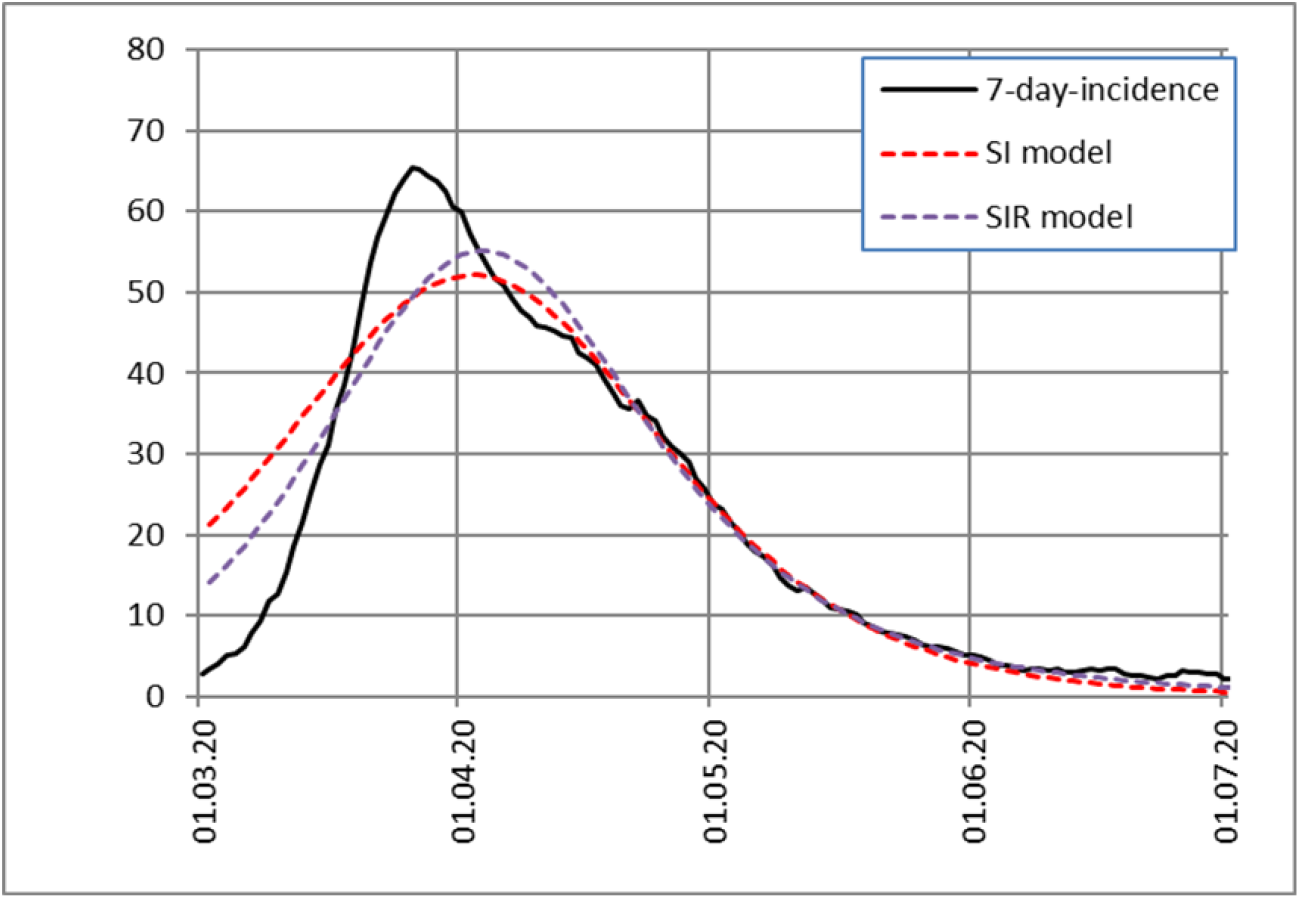
Asymmetrical wave of daily infected in Italy until the end of June 2020.

### SIR model

To improve our modeling of COVID-19 data, we implemented the SIR model [2], which additionally considers the number of resistant, removed, or recovered people R(t):

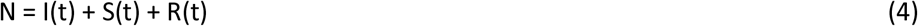

A numerical iterative solution results in 3 equations,

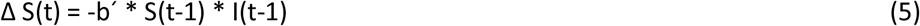

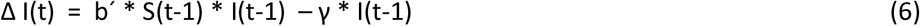

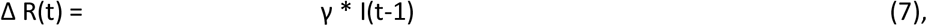

where b’ = b/N is the infection rate and γ (gamma) is the recovery rate. If gamma equals zero, the model is equivalent to the SI model.

Fig. 2 shows the data for new infections in Italy at the end of June 2020. The data were approximated using the SI and SIR models. As expected, the SI model did not agree well with the data. The SIR model leads to an improvement, but still does not fit the data well. Only when we added two SIR model waves did we obtain a very good approximation to the real course in Fig. 3, where we scaled the cases to “per million persons” (“pmp”):

**Fig. 3.**
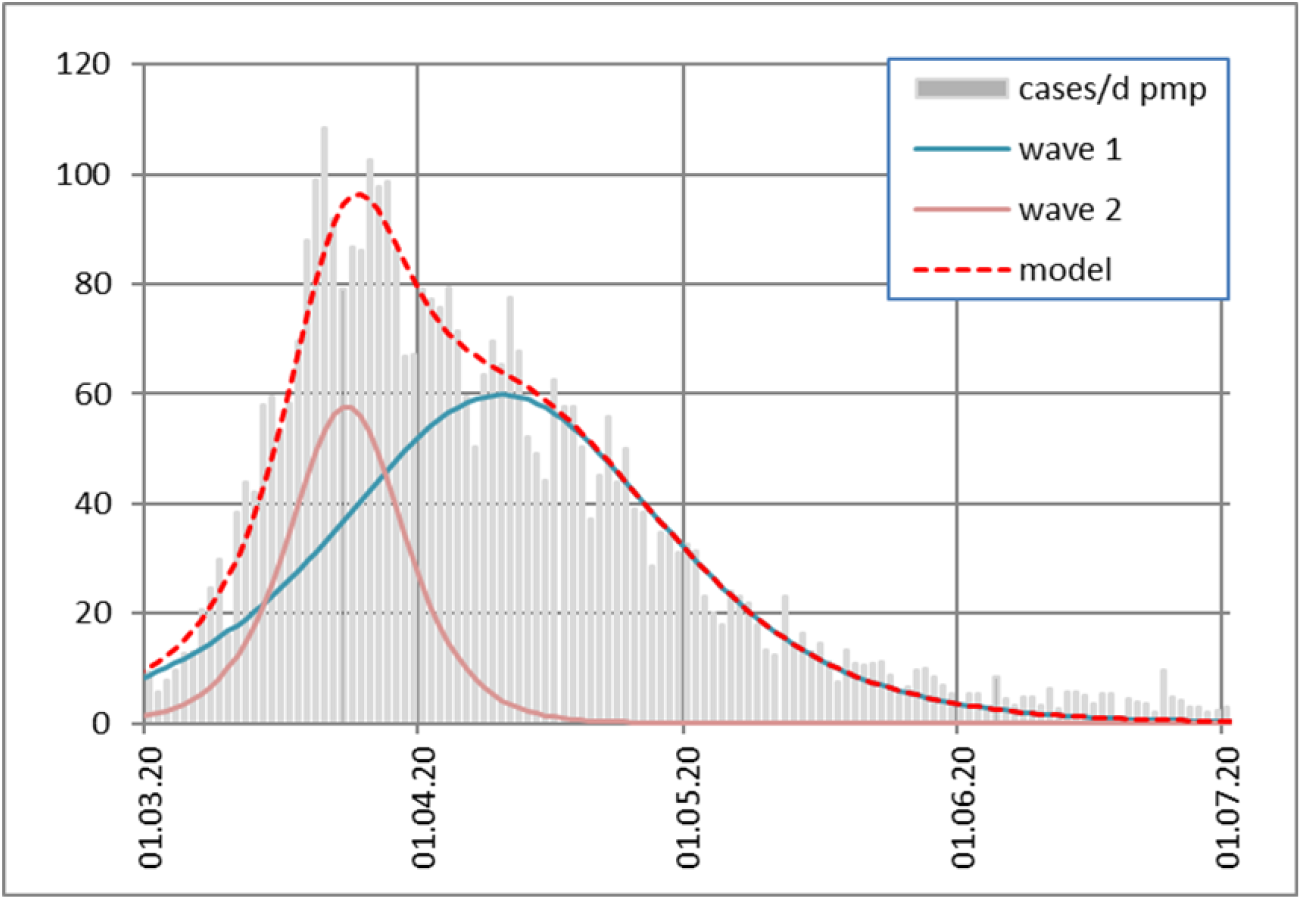
Daily infected in Italy with two model waves

The two different waves may be caused by

- local outbreaks in certain groups or regions
- different test measures
- different preventive measures
- another virus type etc.

Accordingly, we switched to the model with multiple SIR waves in the second wave of COVID-19 to obtain better approximations.

### India as an example of multiple model waves

When we took India as an example for approximating the real course using multiple SIR waves, we also obtained a very good approximation result (Fig. 4):

**Fig. 4.**
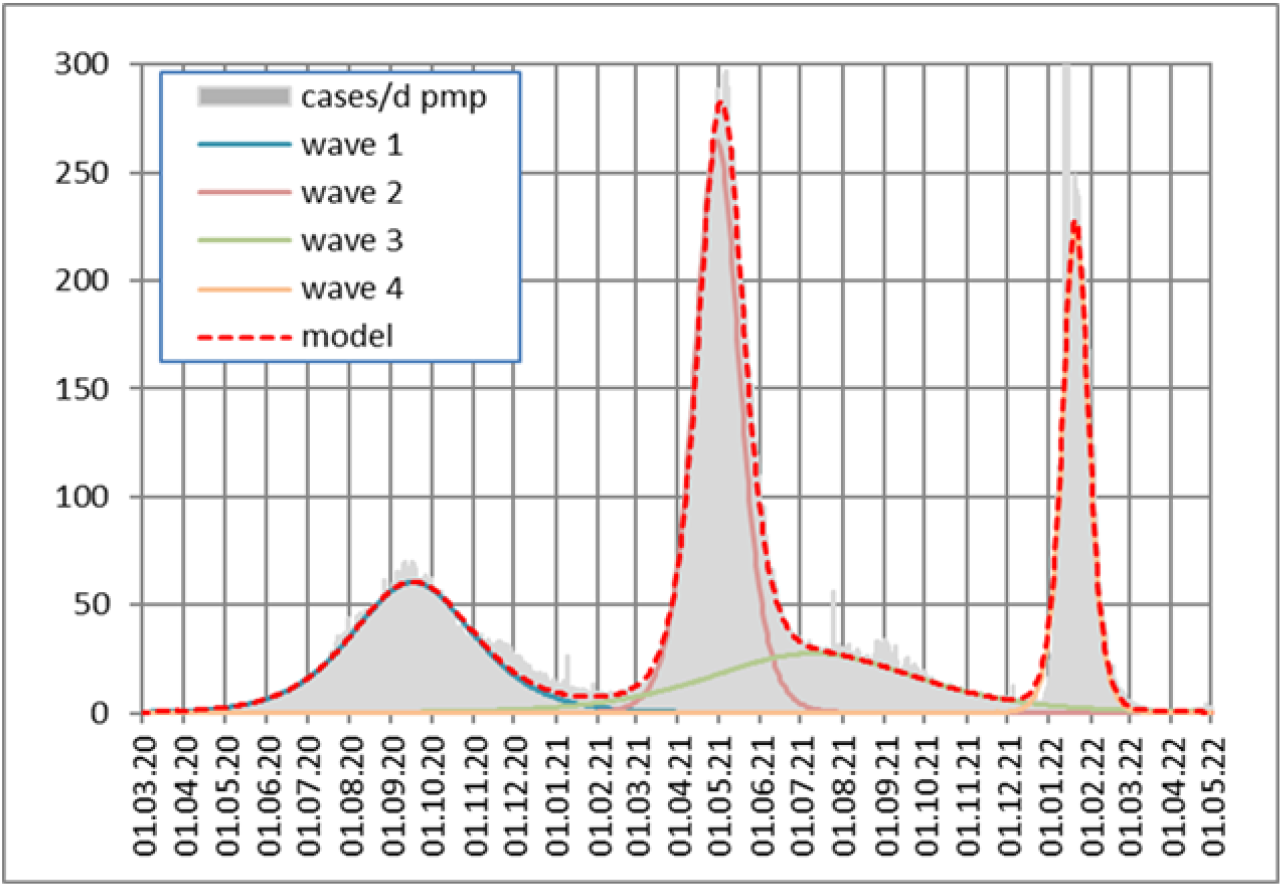
Daily infected in India

However, when we examine the parameters of the iteratively calculated SIR model waves, the SIR parameters are not plausible (Table I).

**Table I.** Modeled SIR waves in India (start day 0 is February 21, 2020)

| wave # | infection rate $b'$ | infected persons pmp per wave, N | start day of next wave | gamma |
| --- | --- | --- | --- | --- |
| 1 | 6 % | 9 383 | 0 | 0.02 |
| 2 | 13 % | 22 034 | 345 | 0.20 |
| 3 | 14 % | **53 063** | 357 | **0.73** |
| 4 | 60 % | **6 566** | 656 | **0.19** |

The start day of a wave was defined as one infected person per million people.

The 3^rd^ flat wave in India does not have an 8 times higher value of infected persons than the 4^th^ wave. This contradiction may be due to the fact, that SIR parameters are nonlinearly interconnected and cannot be separated unambiguously during the iteration process.

As the approximated SIR parameters apparently do not provide insight into a wave, we switched back to the SI models.

When we applied the SI model waves to India instead of SIR waves, we obtained reasonable results. The 3^rd^ and the 4^th^ wave now have similar values, and all infection rates appear to be realistic for each wave (Table II).

**Table II.** Modeled SI waves in India (start day 0 is February 20, 2020)

| wave # | infection rate $b'$ | infected persons pmp per wave, N | start day of next wave | gamma |
| --- | --- | --- | --- | --- |
| 1 | 3 % | 7 319 | 0 | 0 |
| 2 | 9 % | 11 686 | 333 | 0 |
| 3 | 2 % | **5 826** | 34 | 0 |
| 4 | 15 % | **5 821** | 642 | 0 |

### Spain as an example of multiple SI waves

In addition, for more complicated courses, using the SI model waves shows a low deviation between the course and model (Fig. 5, Fig. A12). Fig. 6 shows ten modeled SI waves. The incidence scale in Fig. 5 is 0.7 times lower than the cases per day pmp scale in Fig. 6, because the incidence relates to a sum of 7 days and 100 000 people, while the cases per day relate to one day and 1 000 000 people.

**Fig. 5.**
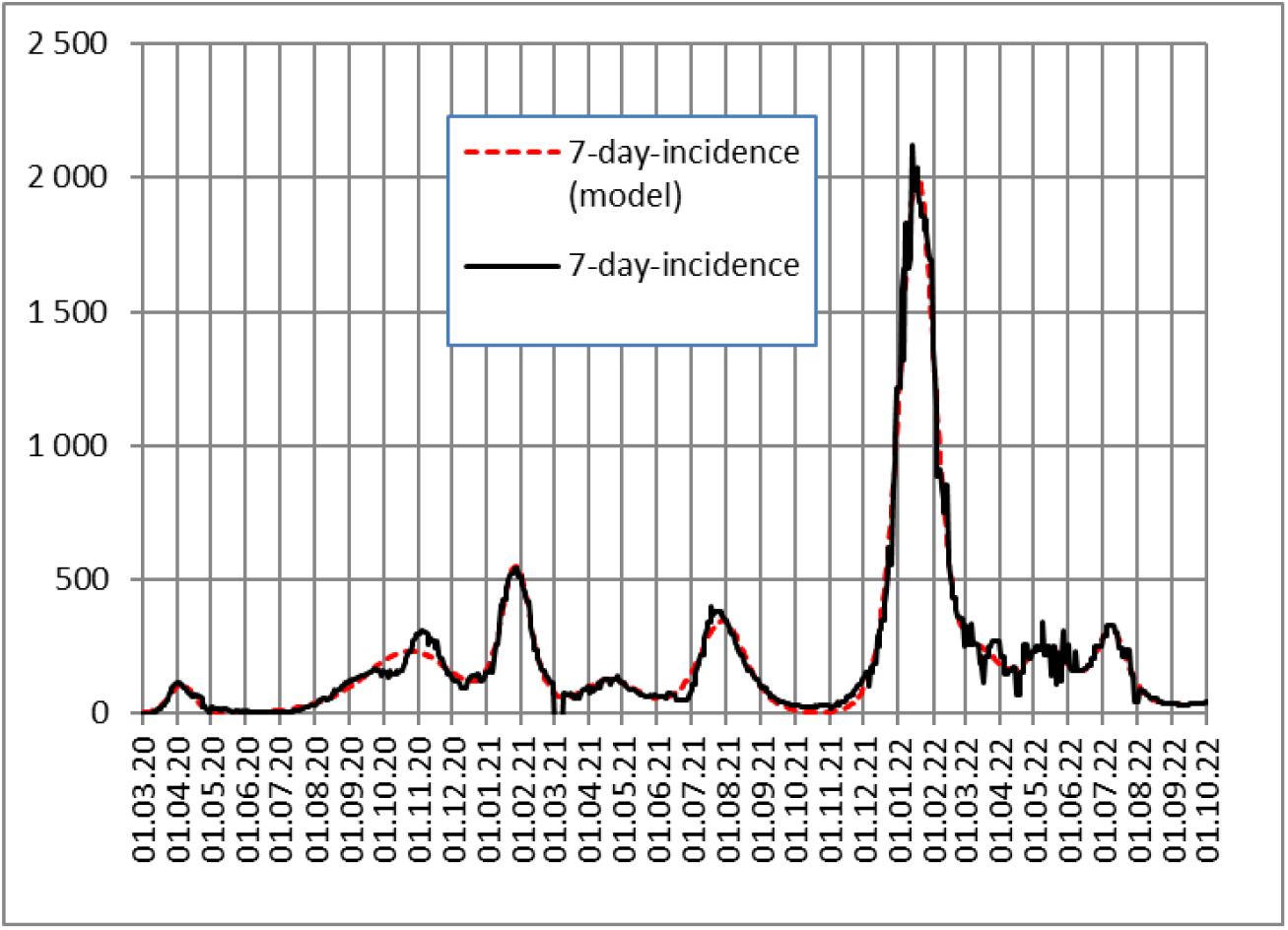
Daily infected in Spain

**Fig. 6.**
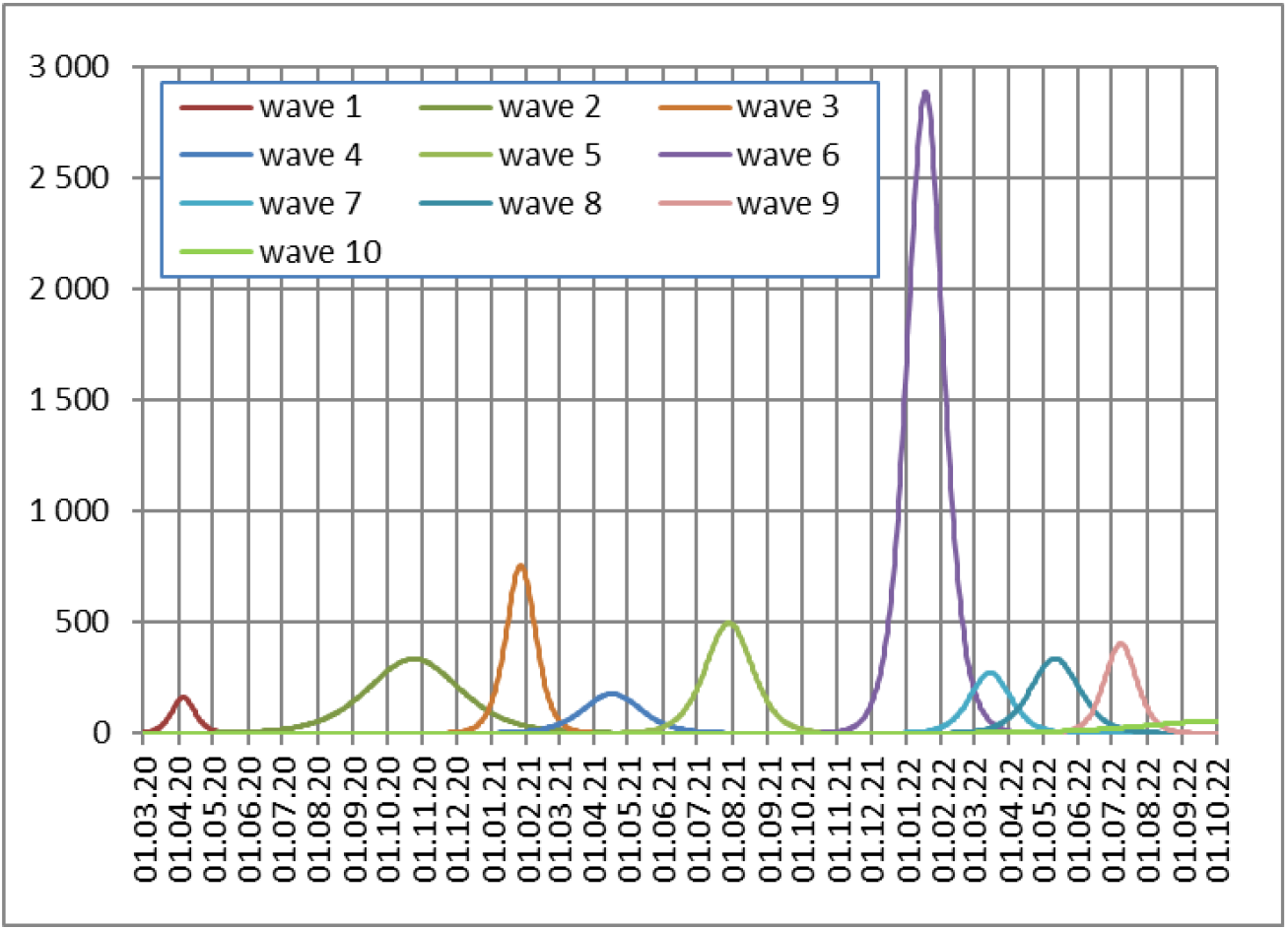
Daily infected in Spain, broken into SI model waves (cases/d pmp)

A new wave is modeled, if the total course approximation is improved.

### Approximation error

Table III lists the relative errors in our model for different countries. The mean daily deviation of our model is given in relation to the mean value of the daily cases.

**Table III.** Approximation error per country

| Country | Mean approximation error 2022 |
| --- | --- |
| AUT Austria | 3.6 % |
| BRA Brazil | 8.3 % |
| ESP Spain | 8.3 % |
| DEU Germany | 2.9 % |
| FRA France | 5.9 % |
| GBR Great Britain | 6.1 % |
| GRC Greece | 3.9 % |
| IND India | 6.7 % |
| ITA Italy | 3.8 % |
| ISR Israel | 7.6 % |
| JPN Japan | 3.1 % |
| NLD Netherlands | 7.4 % |
| SWE Sweden | 11.0 % |
| USA | 4.8 % |
| World | 2.7 % |

The mean relative error of the countries investigated in Table III was approximately 5.8 %.

In 2022, some countries began reporting only once a week. Therefore, the mean approximation error will increase in the future.

### SI waves and virus types

In Germany, the predominant virus types at each time are published by the RKI [6], and we assigned them to SI waves. For some virus types, more than one wave occurred, as indicated by the Roman numbers in Fig. 7, Fig. A04, and in Table A04:

**Fig. 7.**
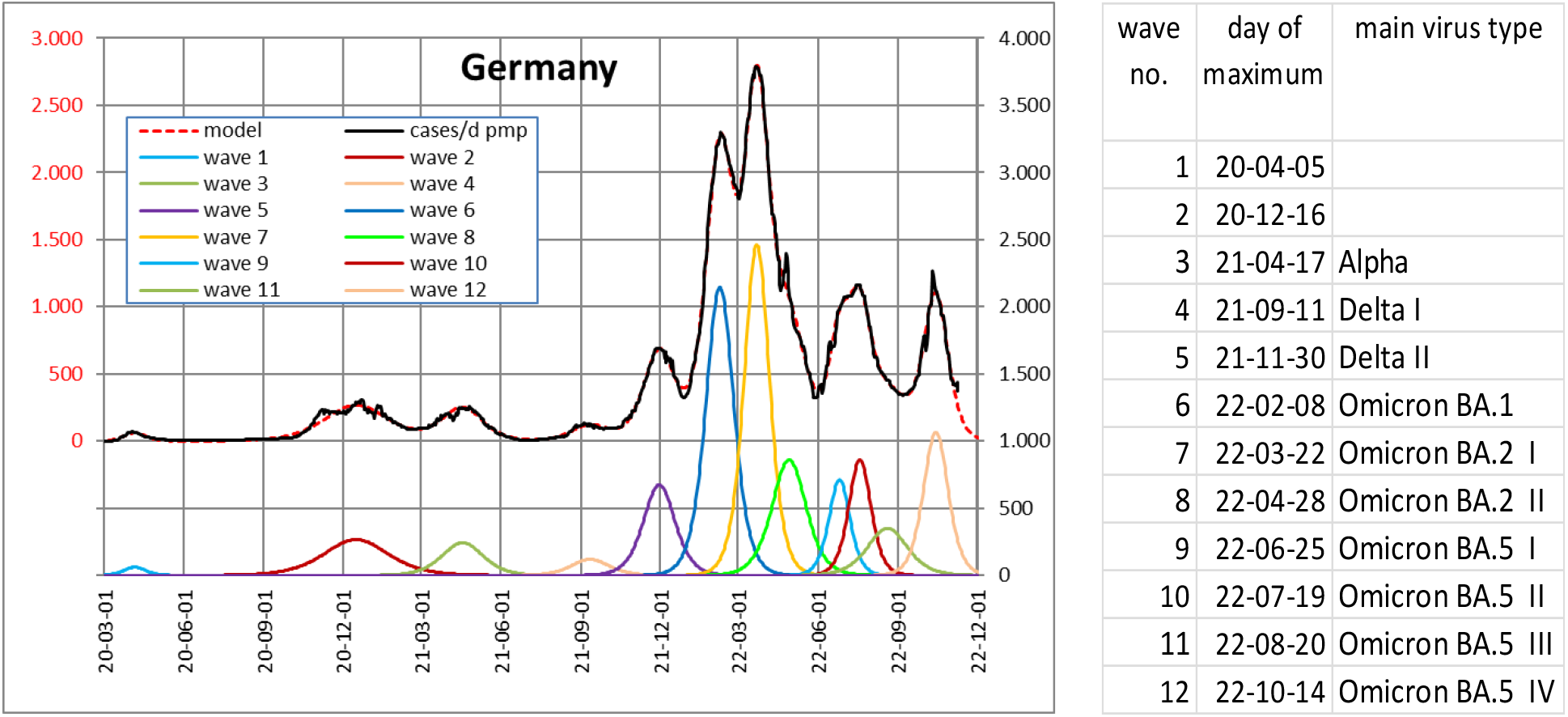
Daily infected in Germany (Updated. Left axis: real course and model. Right axis: model waves)

Even when no virus type data are available, it may be possible to assign the virus types by comparing the waves of different countries during the same time interval.

### SI waves for prediction

The last model wave of a real course may appear to be suitable for predicting the further progression of infections. This may be possible once the wave has reached its maximum in reality. However, at the start of a wave, the slope of an SI function will be nearly independent of the later maximum (Fig. 8). This means that slight variations in the real data result in strong variations in the time point and height of the model maximum.

**Fig. 8.**
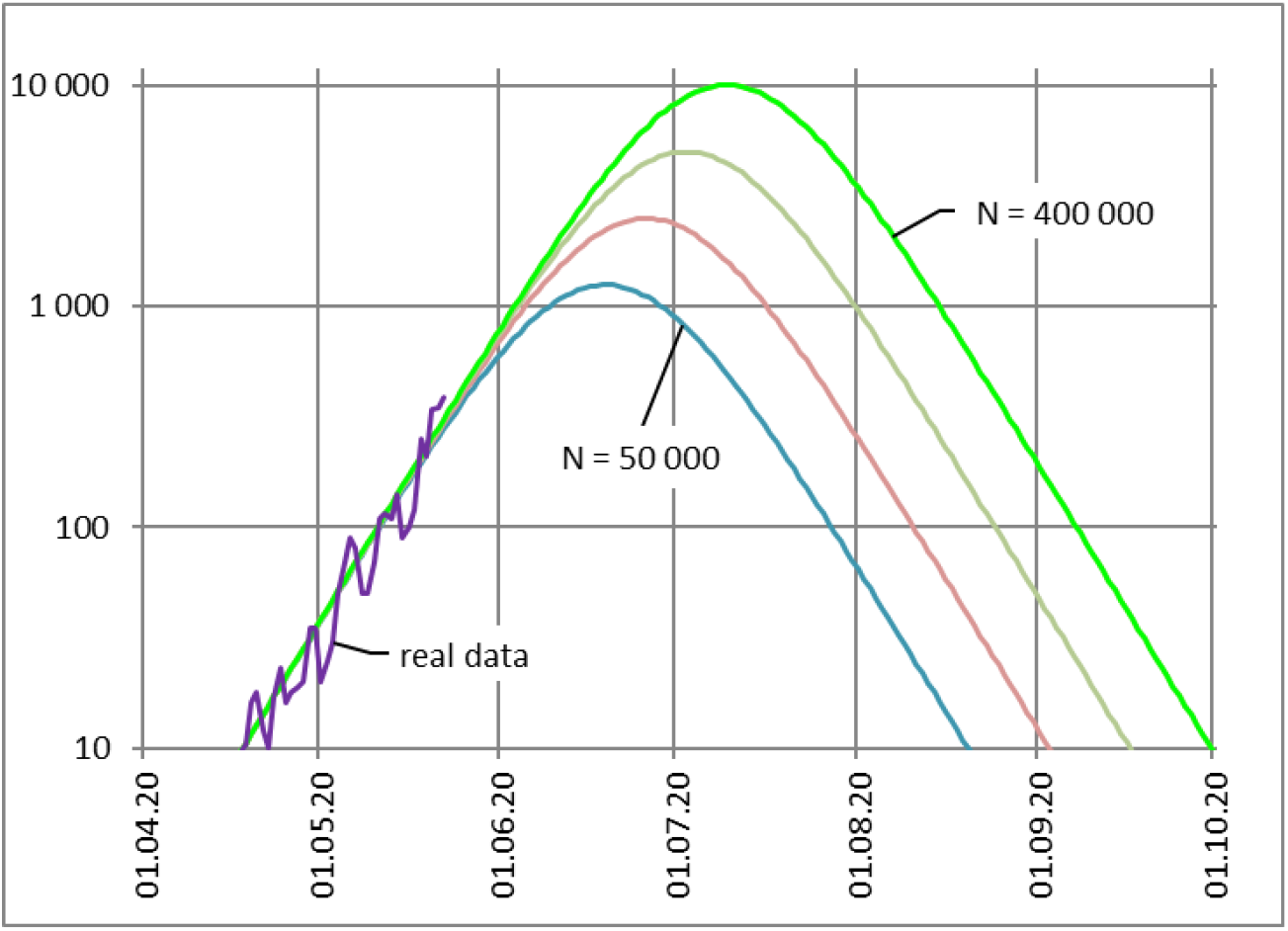
SI waves with different peaks but similar positive slope (logarithmic scale)

We must wait until the slope of the real wave becomes negative to ensure that the maximum has been reached. A positive slope still indicates the minimum number of persons who will be infected until the end of the wave. However, if the negative slope of the real data deviates strongly from the modeled waves, it indicates a new wave. After June 2020, this happened almost every time in all the investigated countries during a negative slope (Fig. A01 to Fig. A14).

### Using SI parameters

SI waves are comparable or sortable by each parameter, including the start day, calculated maximum day, or duration of a wave. Table IV shows an example in which the Omicron BA.1 I waves are sorted by the number N of infected persons.

**Table IV.** Waves of Omicron BA.1 I, sorted by the number N of infected persons.

| Country | day of maximum | Infection rate | infected in wave BA.1 I (pmp) |
| --- | --- | --- | --- |
| FRA | 22-01-24 | 10,7% | 196.562 |
| NLD | 22-02-05 | 10,4% | 174.105 |
| ISR | 22-01-26 | 17,0% | 168.968 |
| AUT | 22-02-06 | 9,8% | 154.388 |
| ESP | 22-01-17 | 9,3% | 121.113 |
| ITA | 22-01-19 | 10,1% | 119.733 |
| SWE | 22-01-28 | 14,8% | 94.485 |
| DEU | 22-02-08 | 9,3% | 91.956 |
| USA | 22-01-17 | 11,7% | 76.324 |
| GBR | 22-01-05 | 15,8% | 52.371 |
| GRC | 22-01-03 | 29,2% | 39.021 |
| BRA | 22-01-30 | 14,8% | 18.238 |
| JPN | 22-02-06 | 13,9% | 17.257 |
| IND | 22-01-24 | 15,2% | 5.808 |

### Special observations

In Great Britain (Fig. 9, Fig. A05), and world data (Fig. 10, Fig. A15), we also detected very long waves that lasted more than 18 months.

**Fig. 9.**
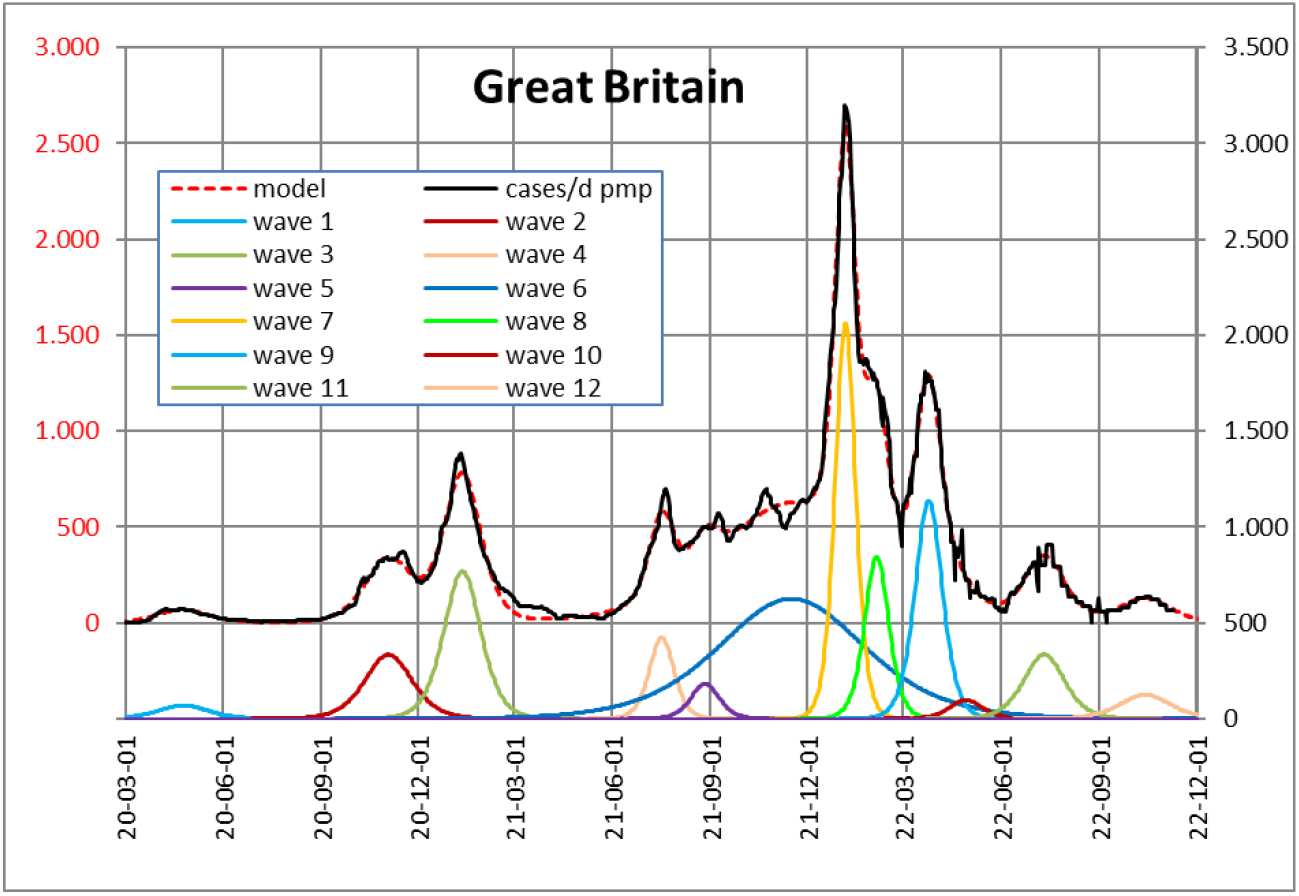
Daily infected in Great Britain, long wave 6 (blue)

**Fig. 10.**
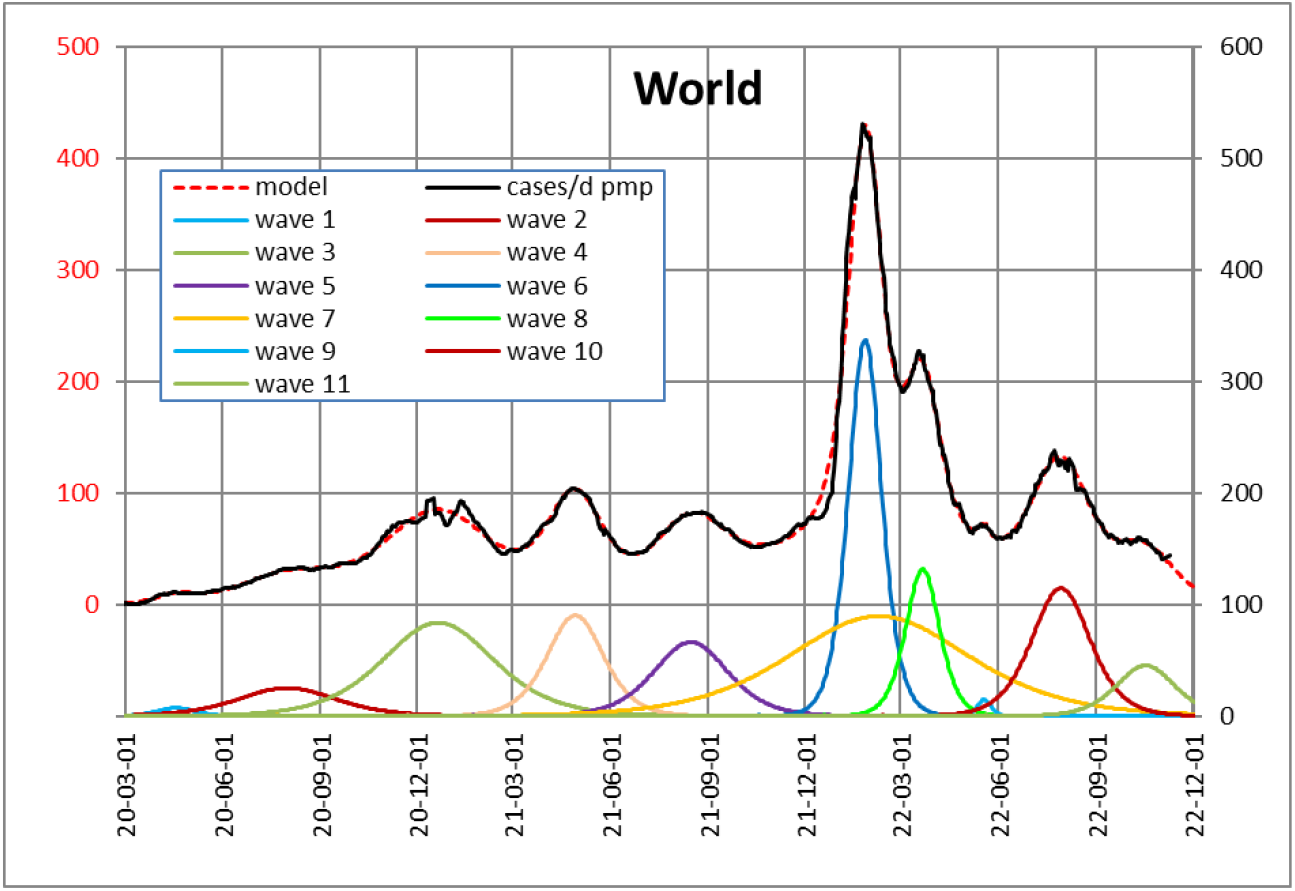
Daily infected in the World, long wave 7

Another interesting effect is that despite lockdowns, vaccinations, or other measures, the total course of the pandemic may be modeled very well by simple symmetrical SI waves. This is obviously due to the fact that lockdowns and other measures only affect the height of a wave, not the shape.

### Publication of the modeling results

Our weekly results are presently used by a local medical COVID-19 committee in the city of Paderborn, Germany.

Link: praxisnetz-pb.de/aktuelles-2/

We used the Excel solver for the iteration of the parameters, allowing for easy use for everyone. Our Excel file also does not include macros. The complete Excel file is available for free download and use. Each of the 190 countries reported by “Our World in Data” is made available by changing the country code within a sheet.

Link: www.janssenplan.de („ activities “)

## Conclusions

The complete COVID-19 course of 14 countries was modeled well by approximately six to 16 SI waves until November 2022. Asymmetrical waves of cases per day were modelled using two or more symmetrical waves. While SIR wave parameters are ambiguous and provide no insight into a wave, we used SI wave parameters that are unambiguous and easy to assign. They are comparable to each other and between countries. The parameters may be correlated to virus types and preventive measures if known in each country. New waves are detectable early, as they show an early deviation from the actual model wave. The last wave of the wave sequence will indicate the progression of the pandemic.

All wave data and the underlying Excel file, including a help sheet, have been published and are free for public use.

## Data Availability

All data produced are available online at
http://www.janssenplan.de/2.html

## Annex

In each Fig. A01–A15 in this annex, the course of infections, the model of the course, and the model waves are shown in a common figure. For better visibility, the waves are shown with a small distance to the real and the model course. In addition, the wave parameters are listed in Tables A01–A15. The start day is omitted if it is outside the range of the figure.

If the input data have been extremely implausible, they have been replaced by the mean of the neighboring values.

**Fig. A01.**
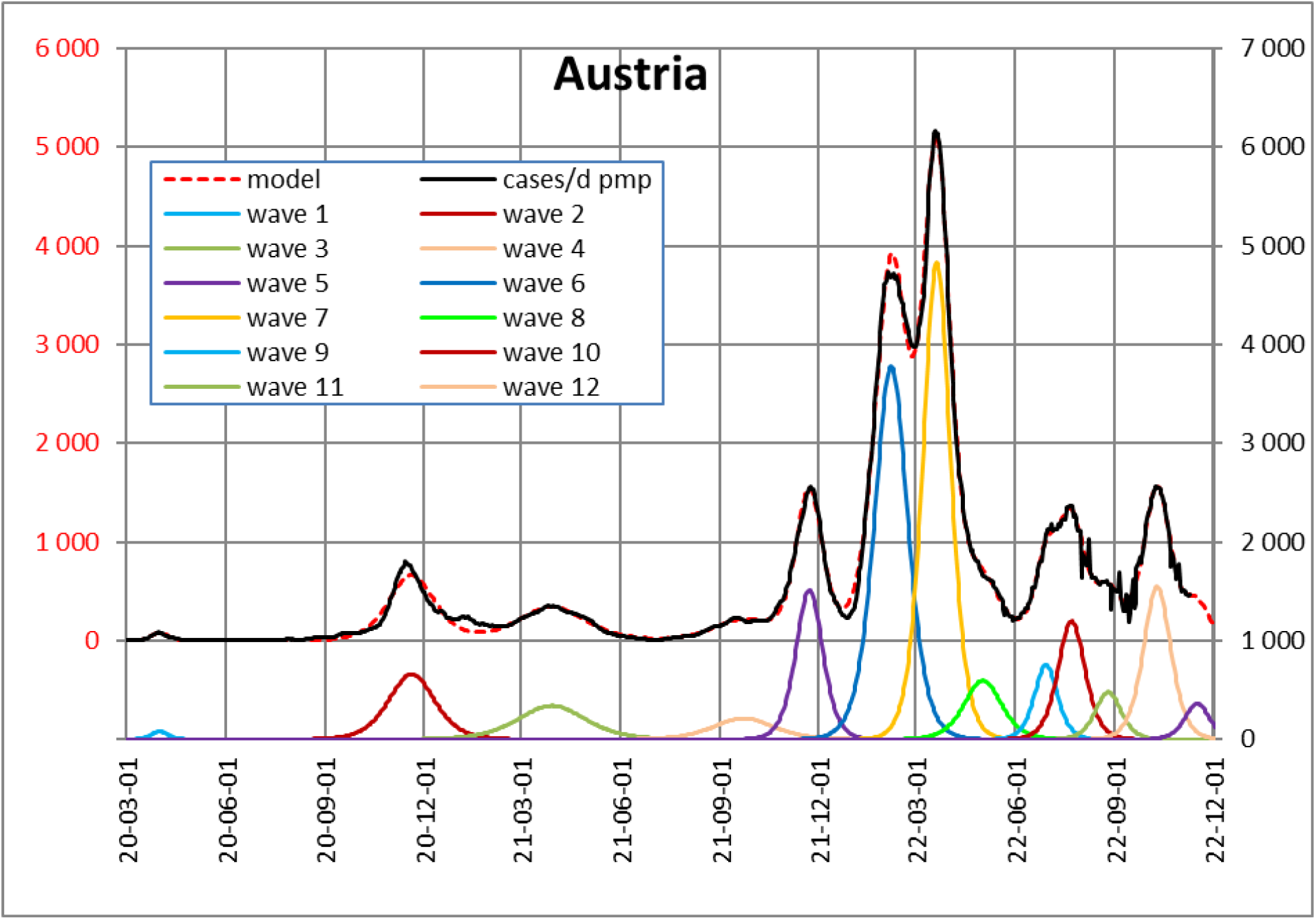
Austria, real course and models

**Table A01.**
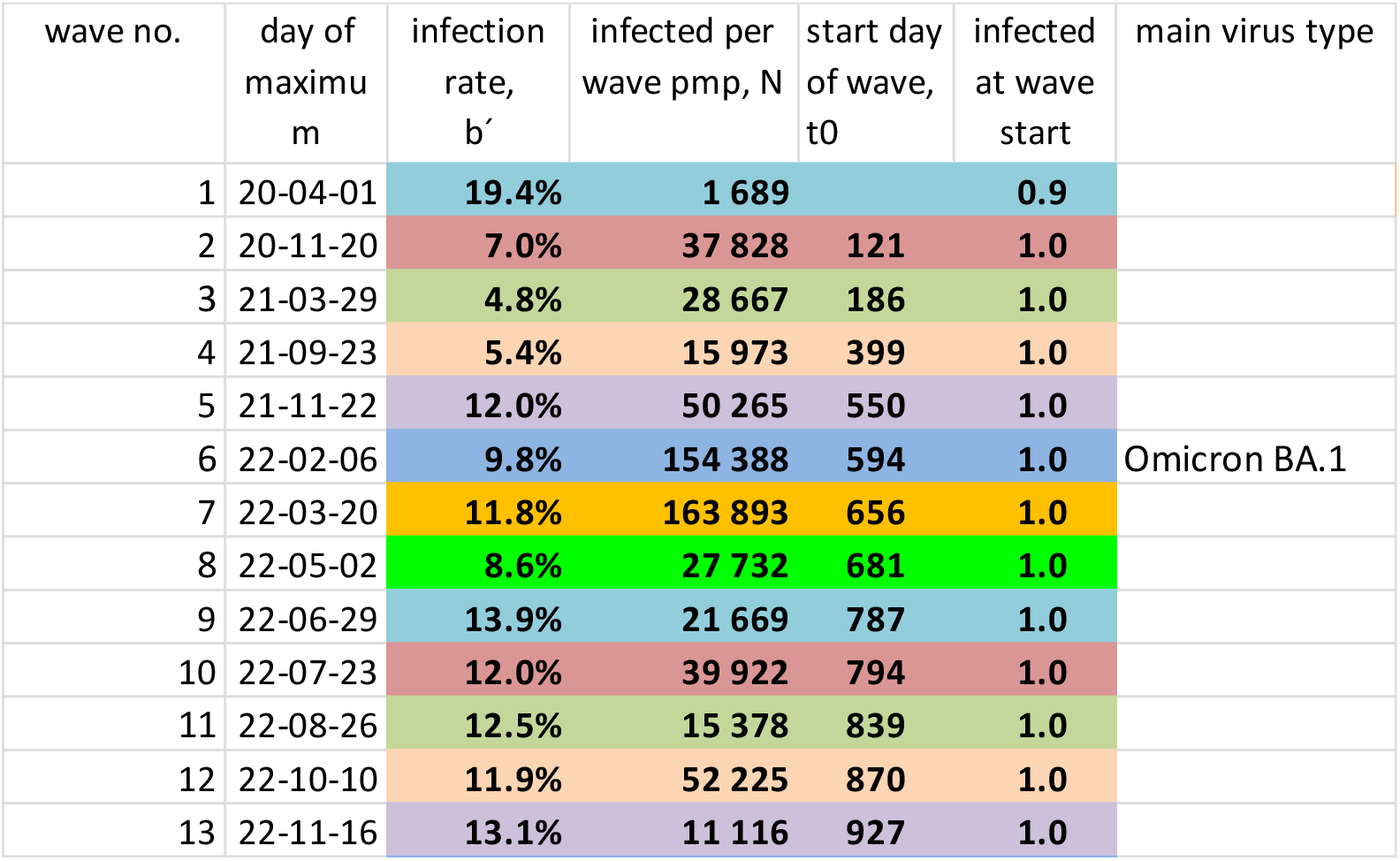
Austria, SI parameters

**Fig. A02.**
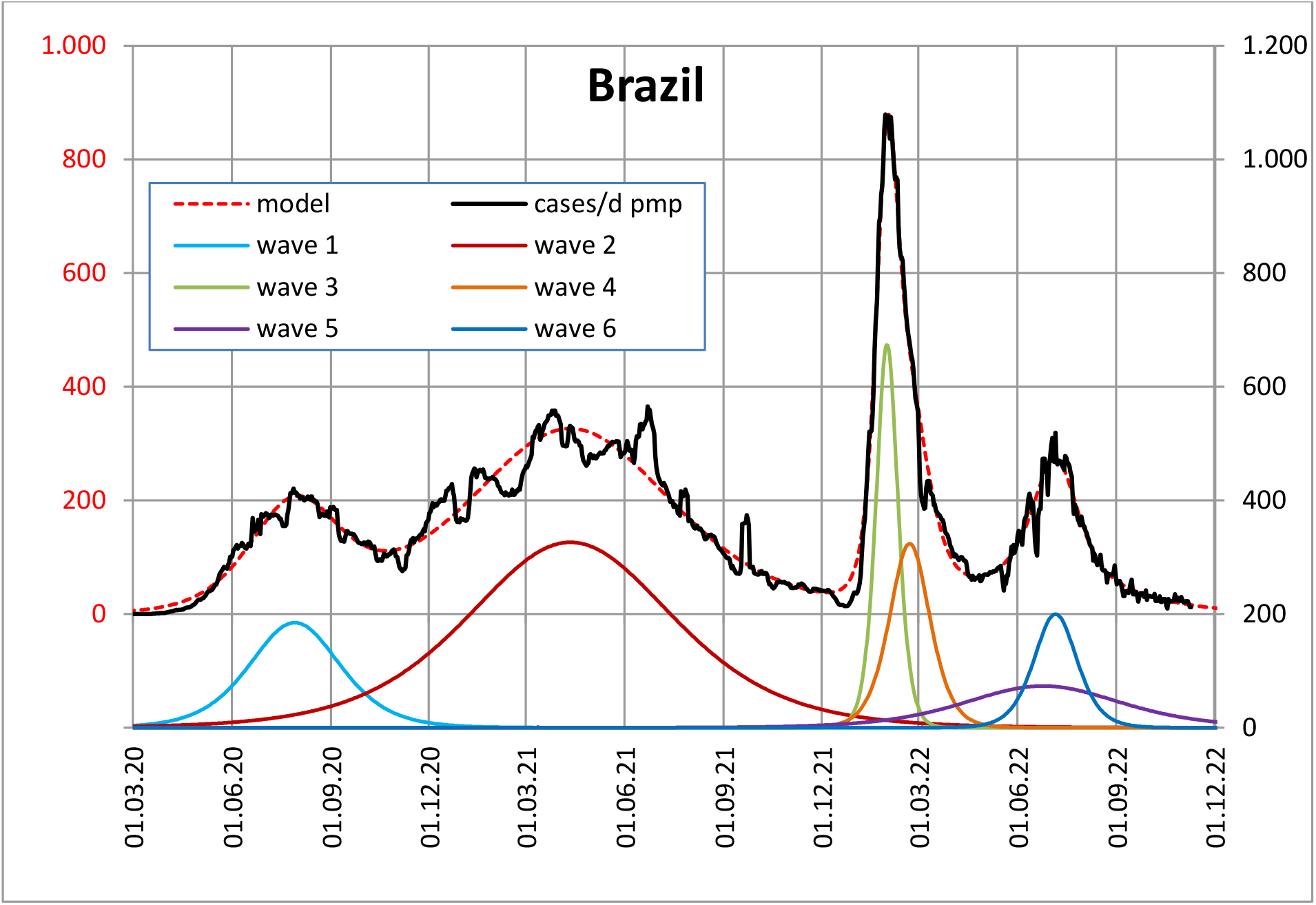
Brazil, real course and models

**Table A02.** Brazil, SI parameters

| wave no. | day of maximum | infection rate, $b'$ | infected per wave pmp, N | start day of wave, $t_0$ | infected at wave start | main virus type |
| --- | --- | --- | --- | --- | --- | --- |
| 1 | 20-07-29 | 3.6% | 20 811 |  | 74.7 |  |
| 2 | 21-04-11 | 1.6% | 83 378 |  | 126.6 |  |
| 3 | 22-01-30 | 14.8% | 18 238 | 643 | 1.0 | Omicron BA.1 |
| 4 | 22-02-21 | 7.9% | 16 303 | 608 | 1.0 |  |
| 5 | 22-06-24 | 2.0% | 14 367 | 385 | 1.0 |  |
| 6 | 22-07-06 | 7.5% | 10 693 | 741 | 1.0 |  |

**Fig. A03.**
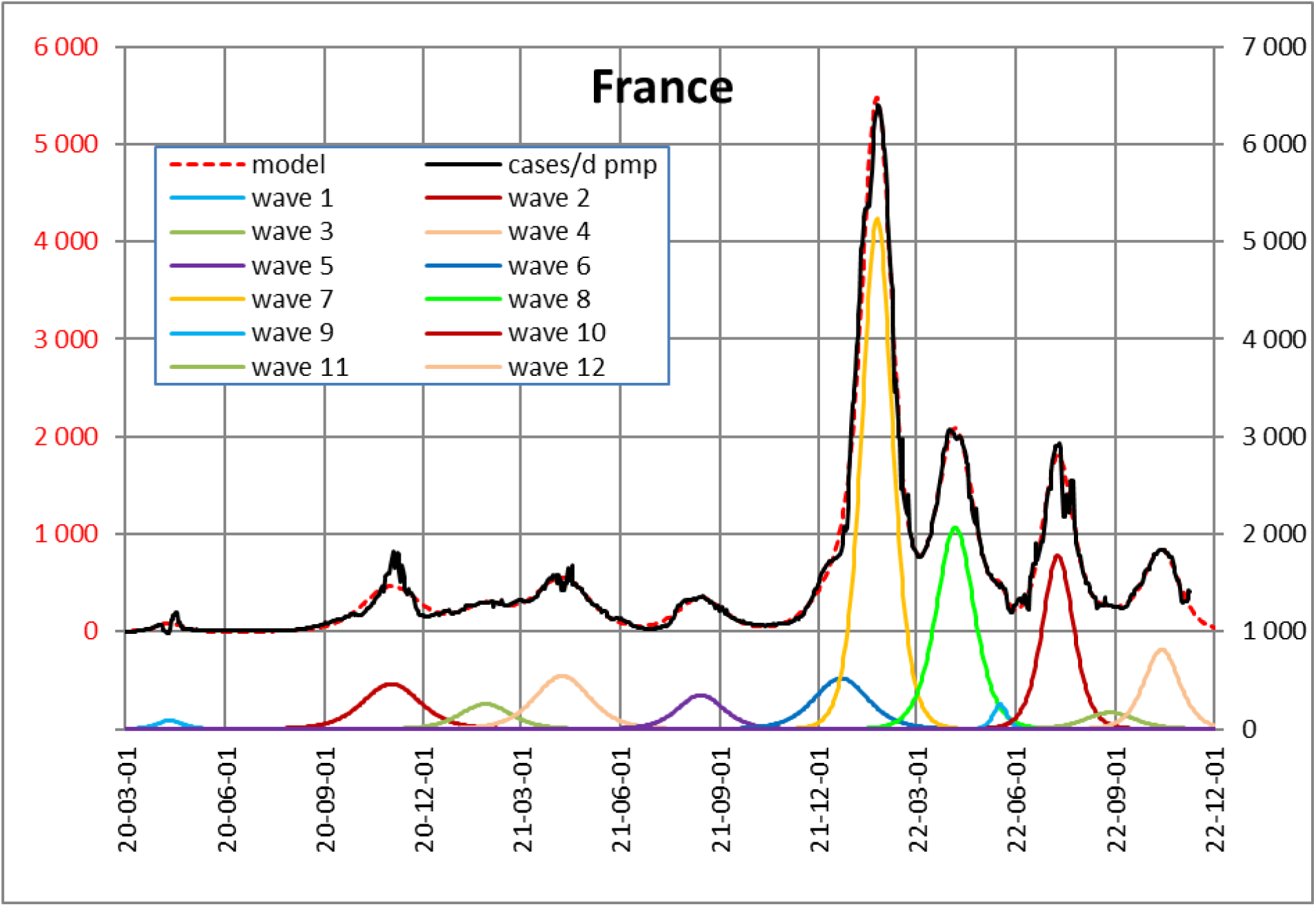
France, real course and models

**Table A03.** France, SI parameters

| wave no. | day of maximum | infection rate, $b'$ | infected per wave pmp, N | start day of wave, $t_0$ | infected at wave start | main virus type |
| --- | --- | --- | --- | --- | --- | --- |
| 1 | 20-04-11 | 12.8% | 2 706 | - | 4.8 |  |
| 2 | 20-11-01 | 5.6% | 32 805 | - | 0.0 |  |
| 3 | 21-01-27 | 6.4% | 15 980 | 190 | 1.0 |  |
| 4 | 21-04-08 | 5.9% | 36 771 | 234 | 1.0 |  |
| 5 | 21-08-14 | 7.1% | 19 690 | 400 | 1.0 |  |
| 6 | 21-12-22 | 6.0% | 34 509 | 496 | 1.0 |  |
| 7 | 22-01-24 | 10.7% | 196 562 | 588 | 1.0 | Omicron BA.1 |
| 8 | 22-04-06 | 8.6% | 96 573 | 641 | 1.0 |  |
| 9 | 22-05-18 | 23.9% | 4 221 | 781 | 1.0 |  |
| 10 | 22-07-10 | 10.4% | 68 130 | 763 | 1.0 |  |
| 11 | 22-08-27 | 6.5% | 10 329 | 776 | 1.0 |  |
| 12 | 22-10-14 | 9.2% | 35 533 | 851 | 1.0 |  |

**Fig. A04.**
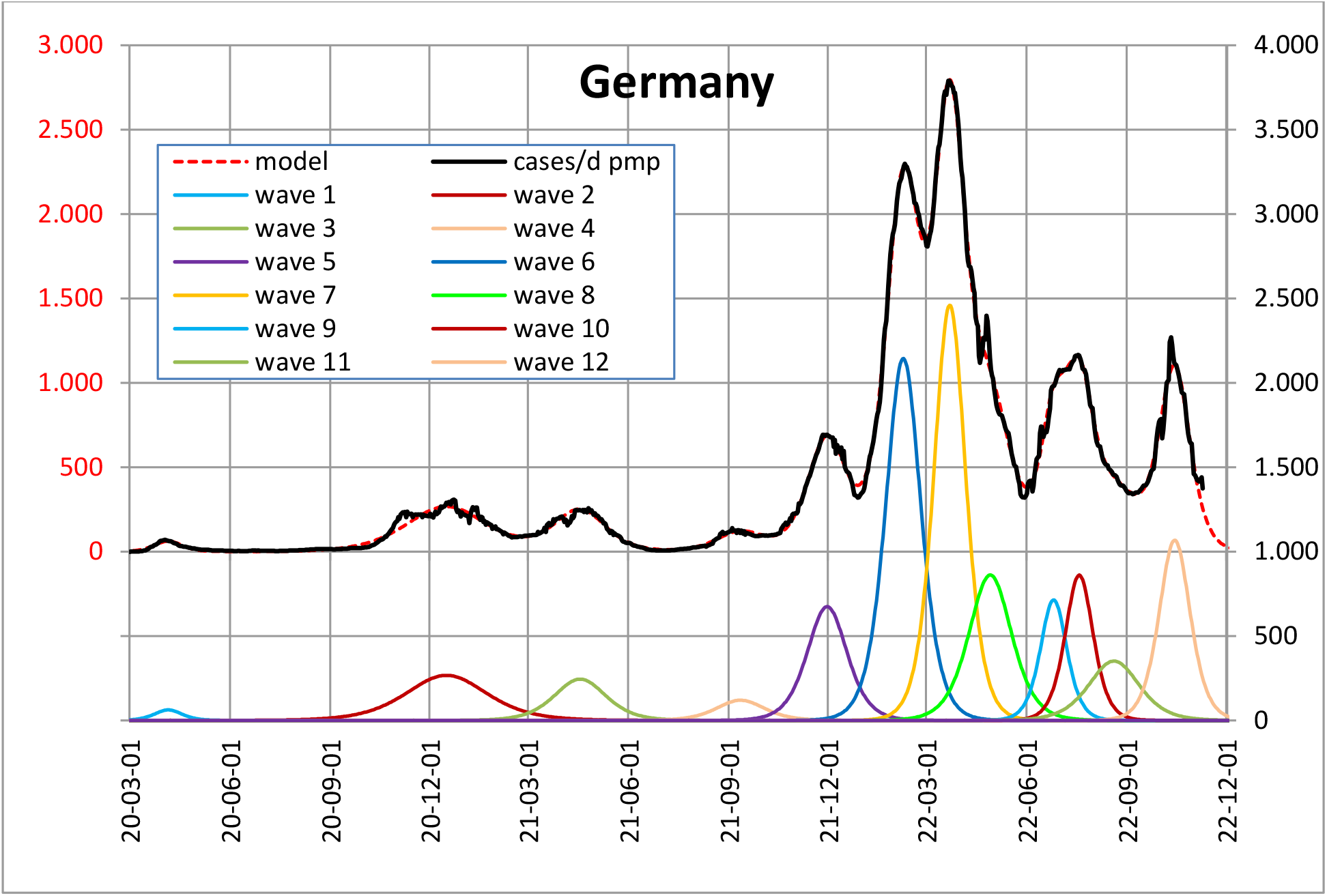
Germany, real course and models

**Table A04.** Germany, SI parameters

| wave no. | day of maximum | infection rate, $b'$ | infected per wave pmp, N | start day of wave, $t_0$ | infected at wave start | main virus type |
| --- | --- | --- | --- | --- | --- | --- |
| 1 | 20-04-05 | 11.8% | 2 124 | - | 12.6 |  |
| 2 | 20-12-16 | 4.0% | 26 582 | 45 | 1.0 |  |
| 3 | 21-04-17 | 6.3% | 15 473 | 268 | 1.0 | Alpha |
| 4 | 21-09-11 | 6.8% | 7 067 | 437 | 1.0 | Delta I |
| 5 | 21-11-30 | 8.2% | 32 900 | 520 | 1.0 | Delta II |
| 6 | 22-02-08 | 9.3% | 91 956 | 595 | 1.0 | Omicron BA.1 |
| 7 | 22-03-22 | 10.3% | 95 695 | 648 | 1.0 | Omicron BA.2 I |
| 8 | 22-04-28 | 7.6% | 45 116 | 657 | 1.0 | Omicron BA.2 II |
| 9 | 22-06-25 | 12.4% | 23 067 | 774 | 1.0 | Omicron BA.5 I |
| 10 | 22-07-19 | 11.7% | 29 380 | 791 | 1.0 | Omicron BA.5 II |
| 11 | 22-08-20 | 6.5% | 21 712 | 756 | 1.0 | Omicron BA.5 III |
| 12 | 22-10-14 | 10.5% | 40 537 | 865 | 1.0 | Omicron BA.5 IV |

Input data are taken from RKI [5], as they are continuously updated also retroactiv. Main virus types are assigned on base of RKI [7].

**Fig. A05.**
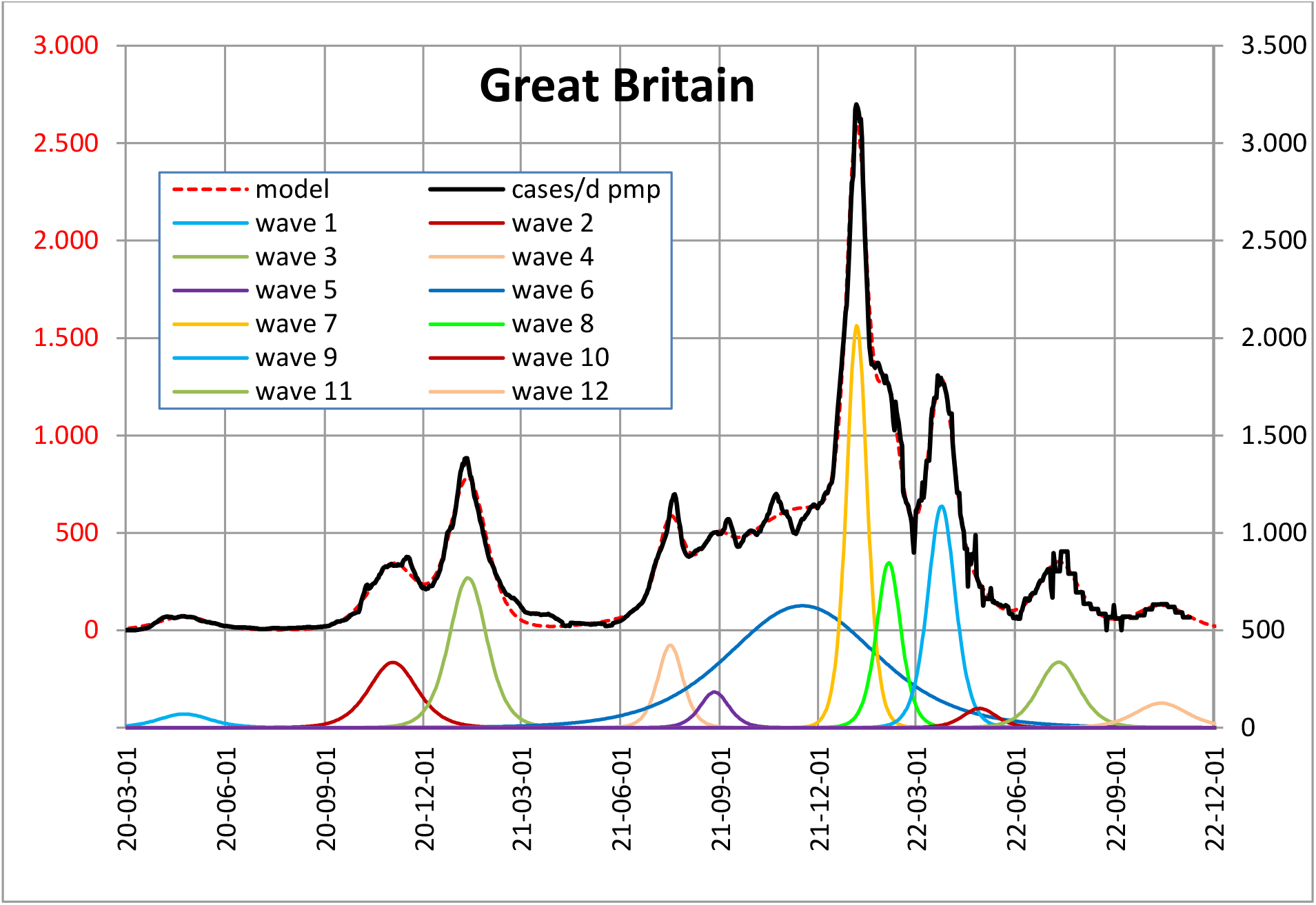
Great Britain, real course and models

**Table A05.**
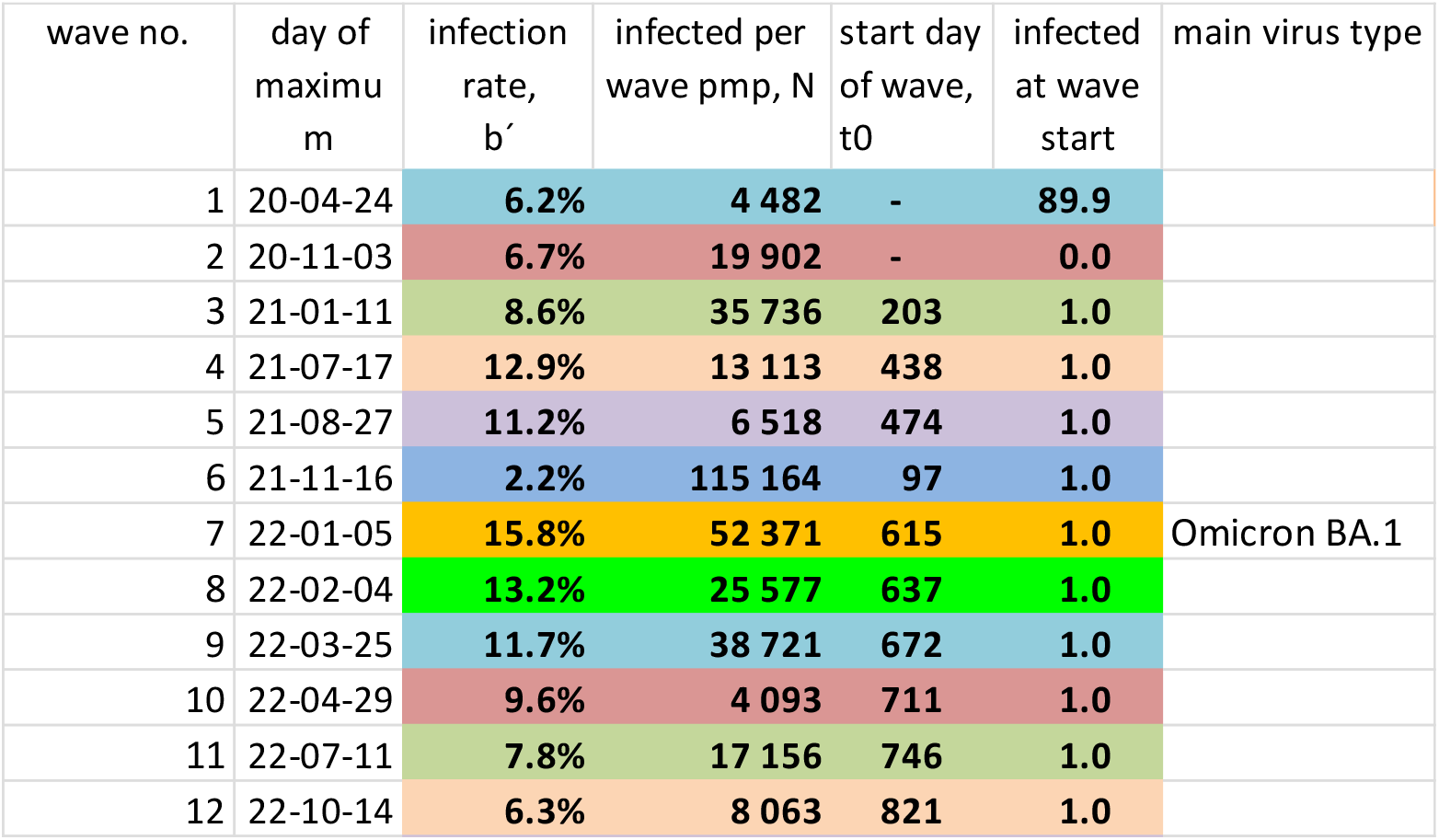
Great Britain, SI parameters

| wave no. | day of maximum | infection rate, $b'$ | infected per wave pmp, N | start day of wave, $t_0$ | infected at wave start | main virus type |
| --- | --- | --- | --- | --- | --- | --- |
| 1 | 20-04-24 | 6.2% | 4 482 | - | 89.9 |  |
| 2 | 20-11-03 | 6.7% | 19 902 | - | 0.0 |  |
| 3 | 21-01-11 | 8.6% | 35 736 | 203 | 1.0 |  |
| 4 | 21-07-17 | 12.9% | 13 113 | 438 | 1.0 |  |
| 5 | 21-08-27 | 11.2% | 6 518 | 474 | 1.0 |  |
| 6 | 21-11-16 | 2.2% | 115 164 | 97 | 1.0 |  |
| 7 | 22-01-05 | 15.8% | 52 371 | 615 | 1.0 | Omicron BA.1 |
| 8 | 22-02-04 | 13.2% | 25 577 | 637 | 1.0 |  |
| 9 | 22-03-25 | 11.7% | 38 721 | 672 | 1.0 |  |
| 10 | 22-04-29 | 9.6% | 4 093 | 711 | 1.0 |  |
| 11 | 22-07-11 | 7.8% | 17 156 | 746 | 1.0 |  |
| 12 | 22-10-14 | 6.3% | 8 063 | 821 | 1.0 |  |

**Fig. A06.**
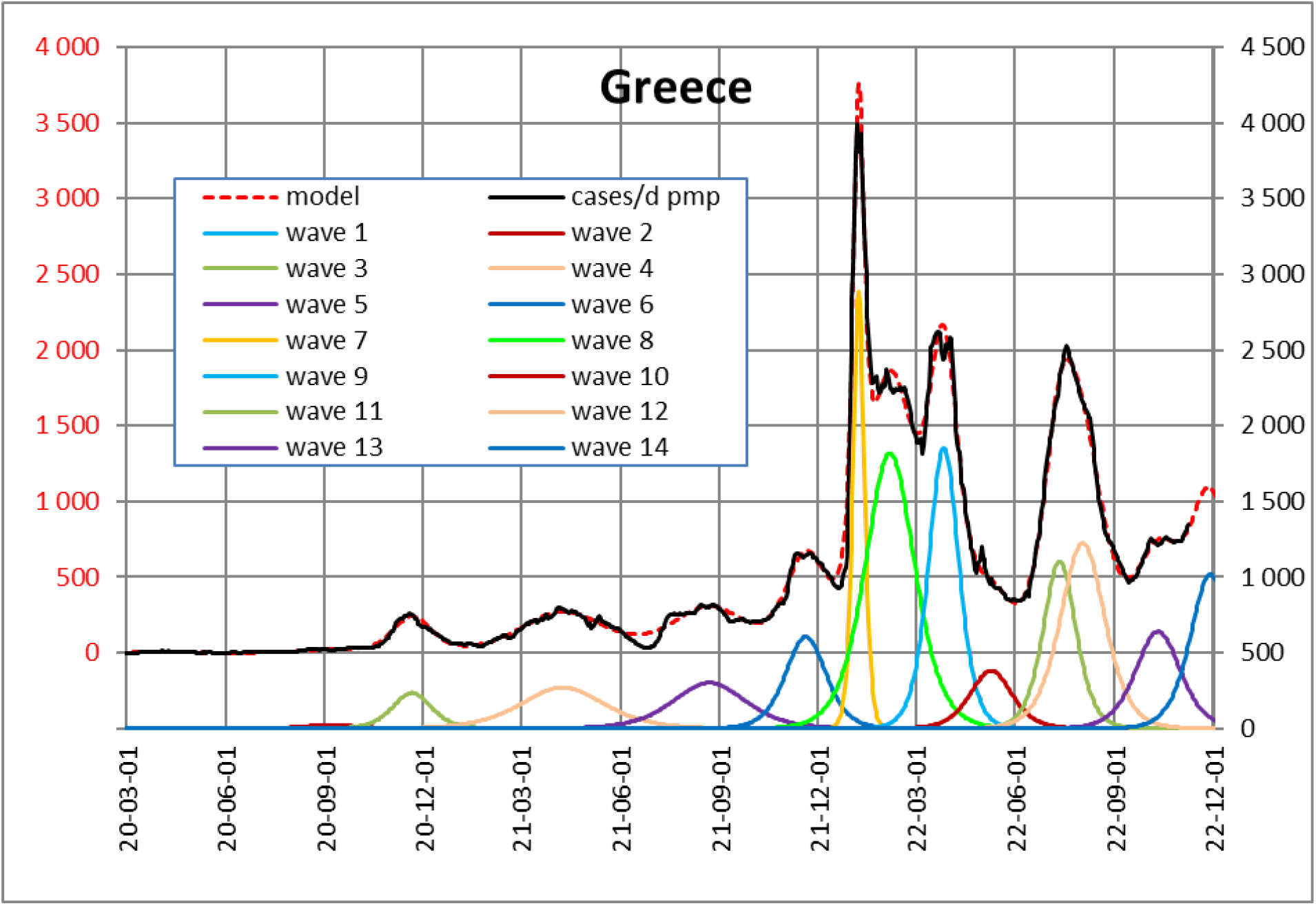
Greece, real course and models

**Table A06.**
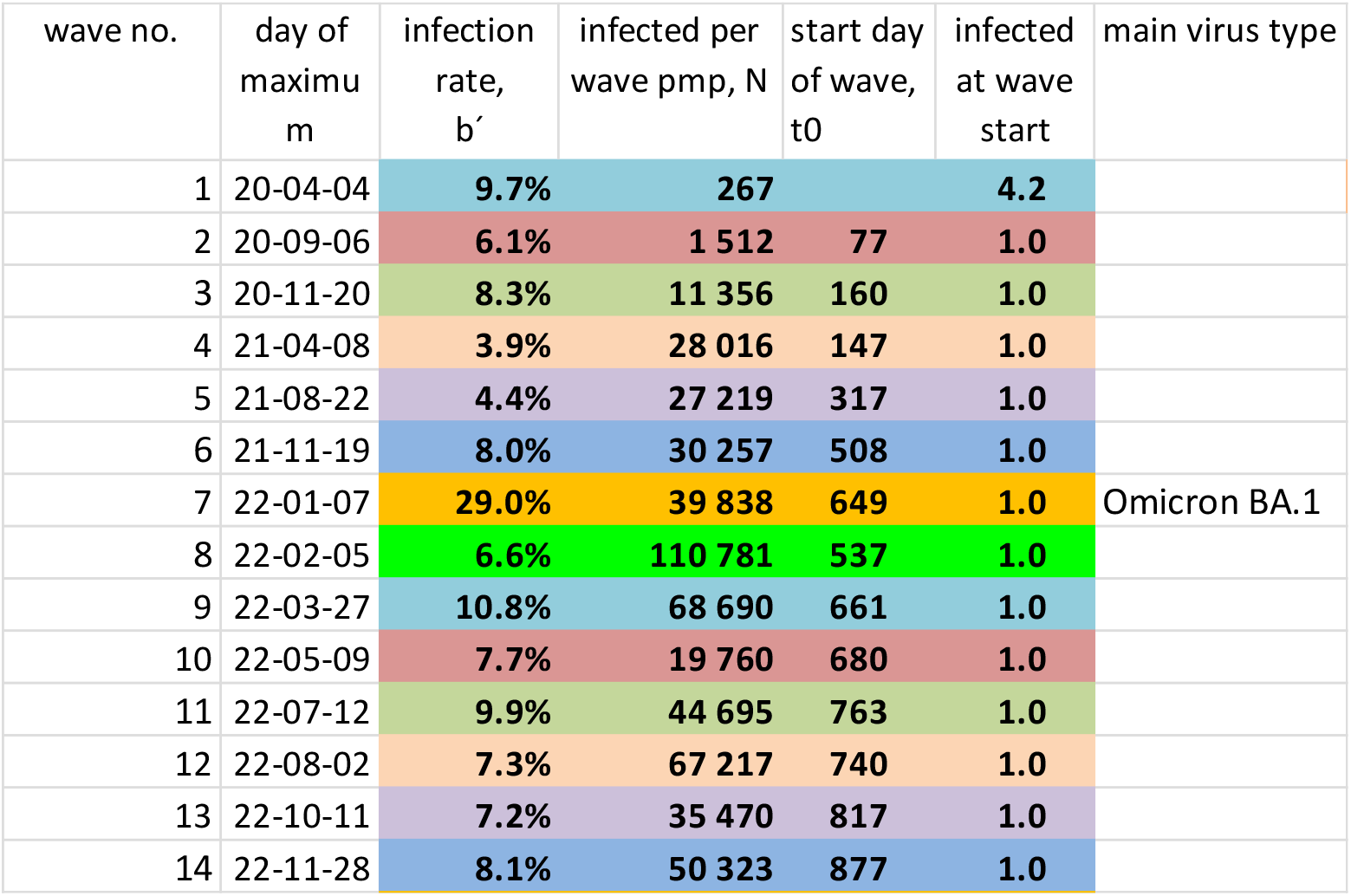
Greece, SI parameters

**Fig. A07.**
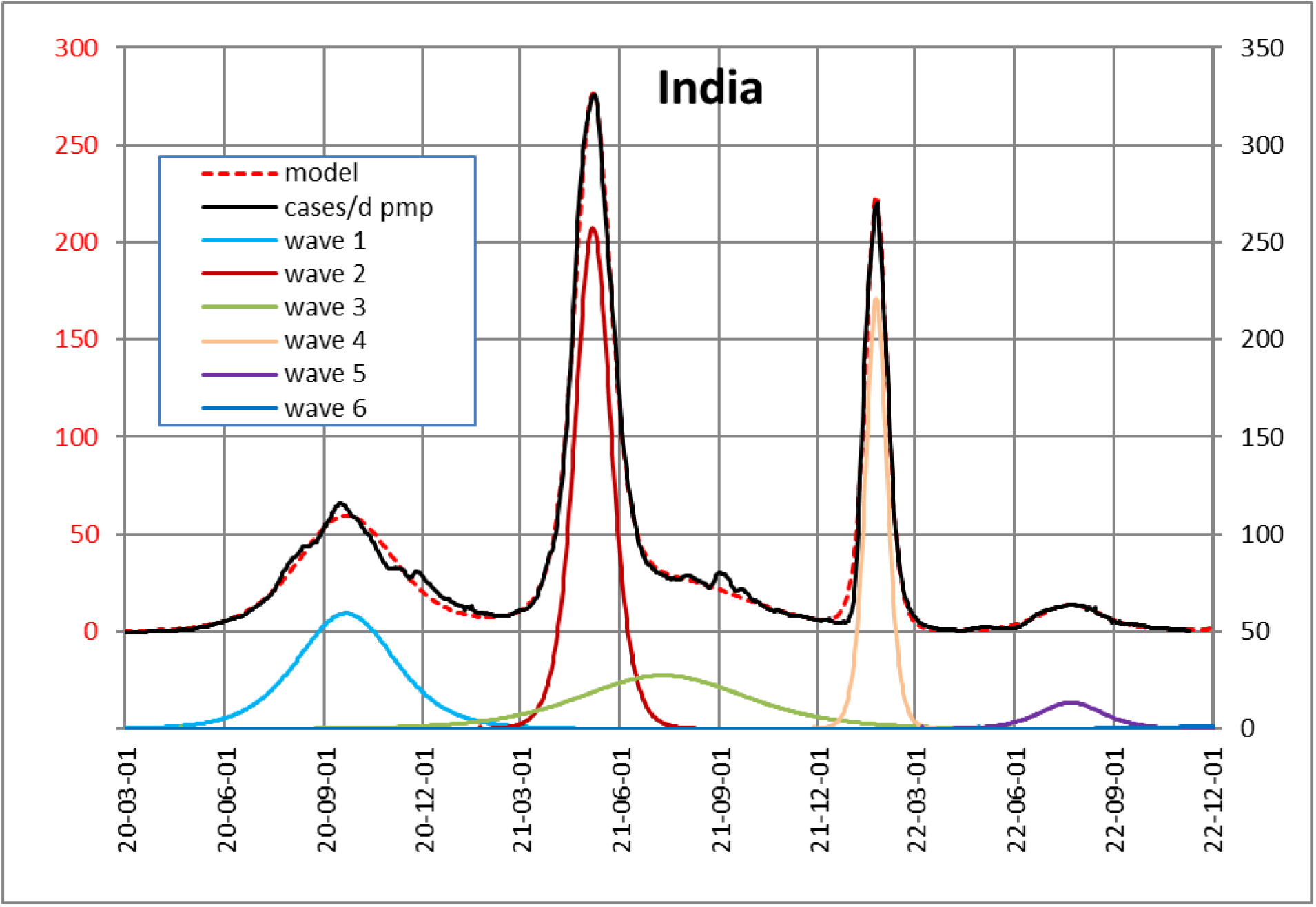
India, real course and models

**Table A07.** India, SI parameters

| wave no. | day of maximum | infection rate, $b'$ | infected per wave pmp, N | start day of wave, $t_0$ | infected at wave start | main virus type |
| --- | --- | --- | --- | --- | --- | --- |
| 1 | 20-09-21 | 3.3% | 7 176 | - | 6.4 |  |
| 2 | 21-05-07 | 9.0% | 11 467 | 336 | 1.0 |  |
| 3 | 21-07-11 | 1.9% | 5 674 | 59 | 1.0 |  |
| 4 | 22-01-24 | 15.2% | 5 808 | 646 | 1.0 | Omicron BA.1 |
| 5 | 22-07-23 | 5.1% | 1 067 | 745 | 1.0 |  |
| 6 | 23-01-26 | 2.2% | 370 | 797 | 1.0 |  |

**Fig. A08.**
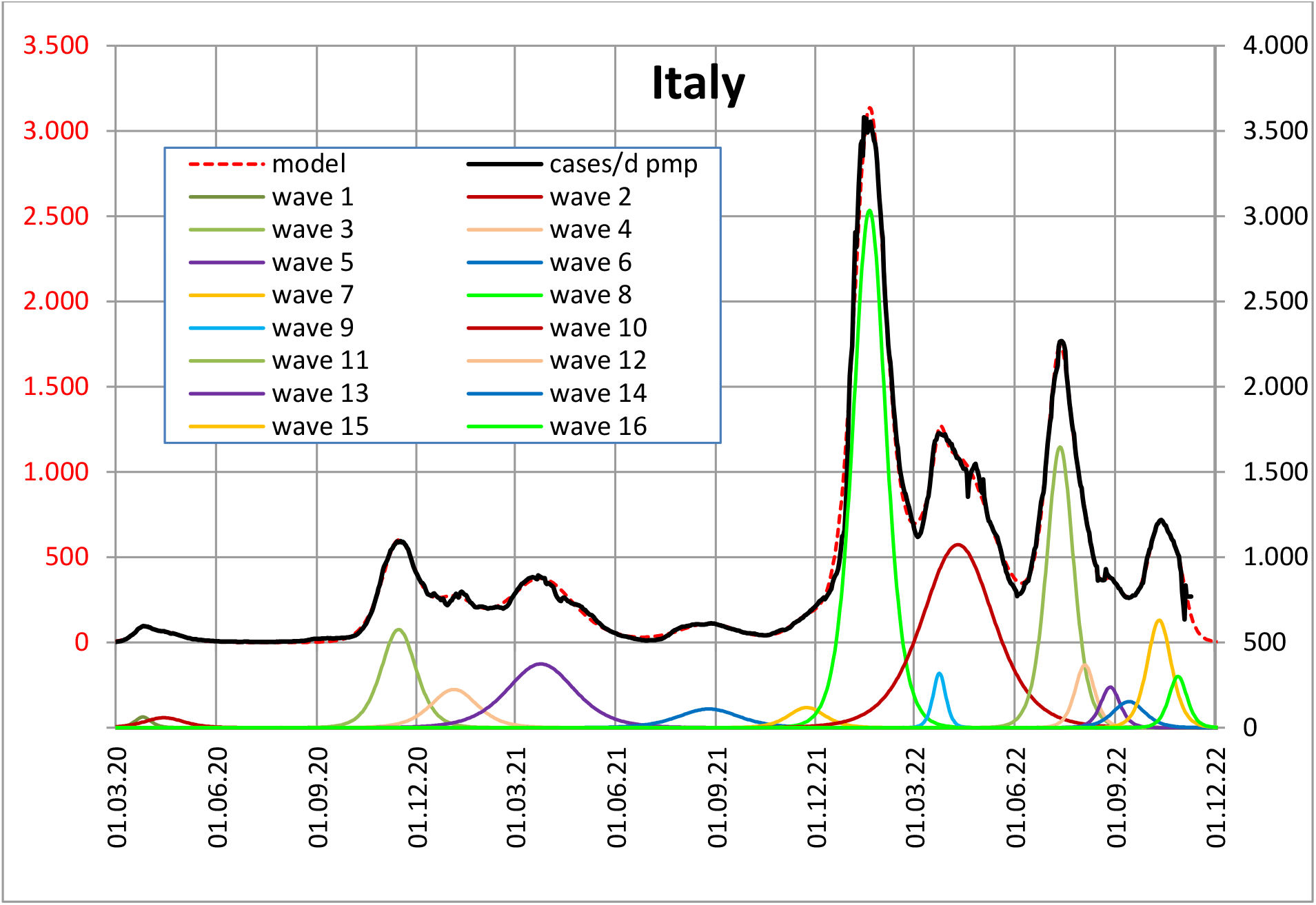
Italy, real course and models

**Table A08.**
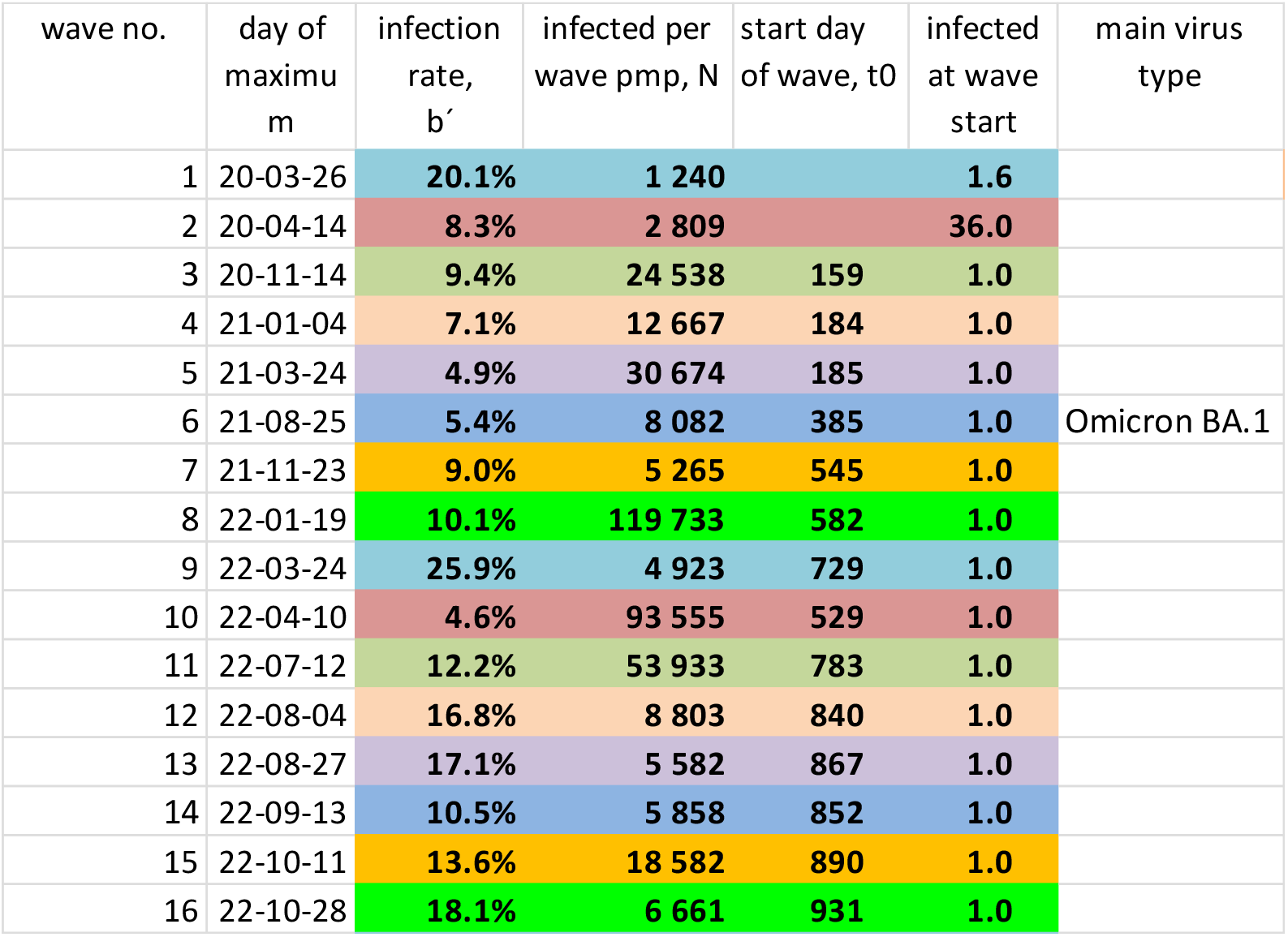
Italy, SI parameters

**Fig. A09.**
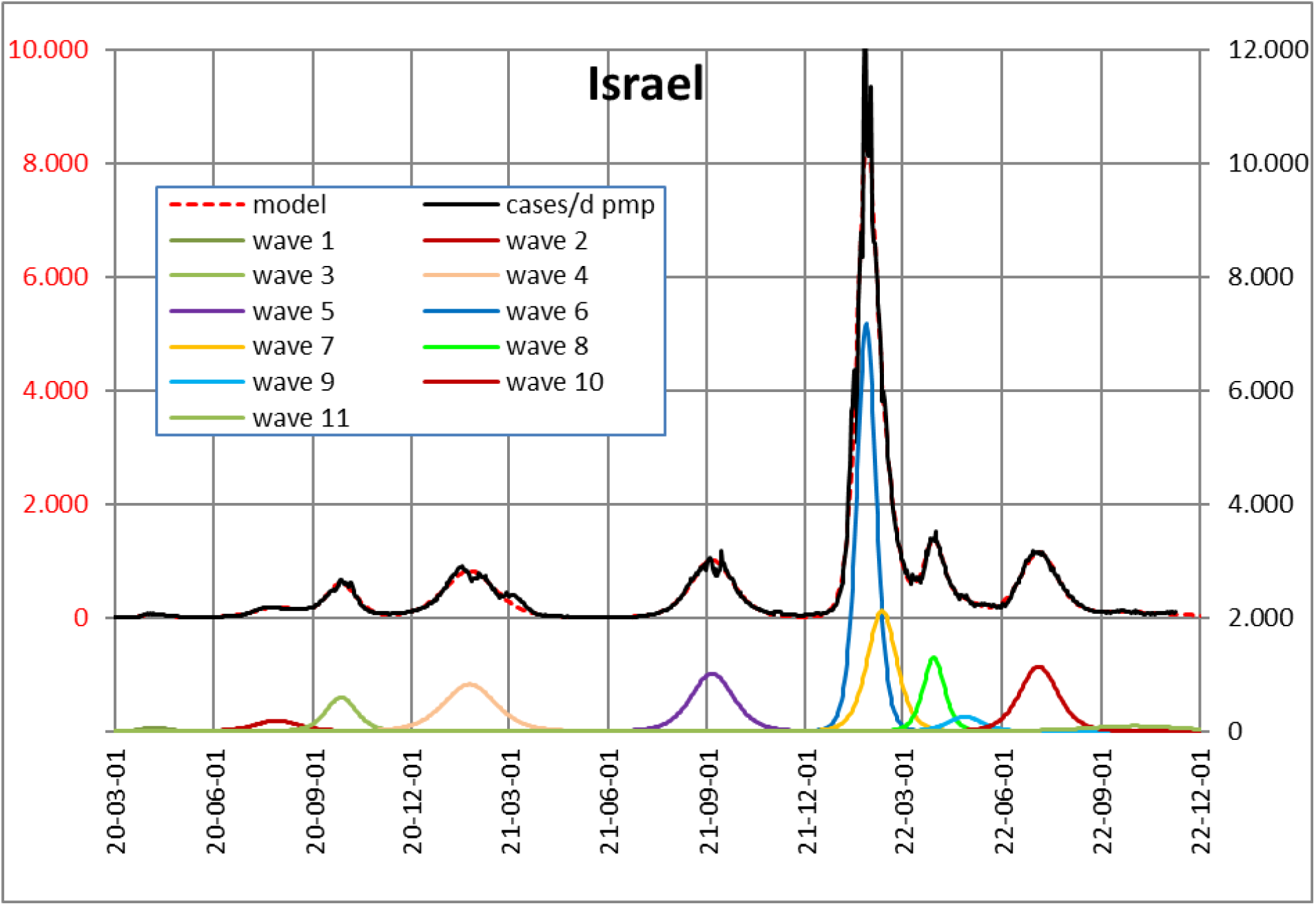
Israel, real course and models

**Table A09.**
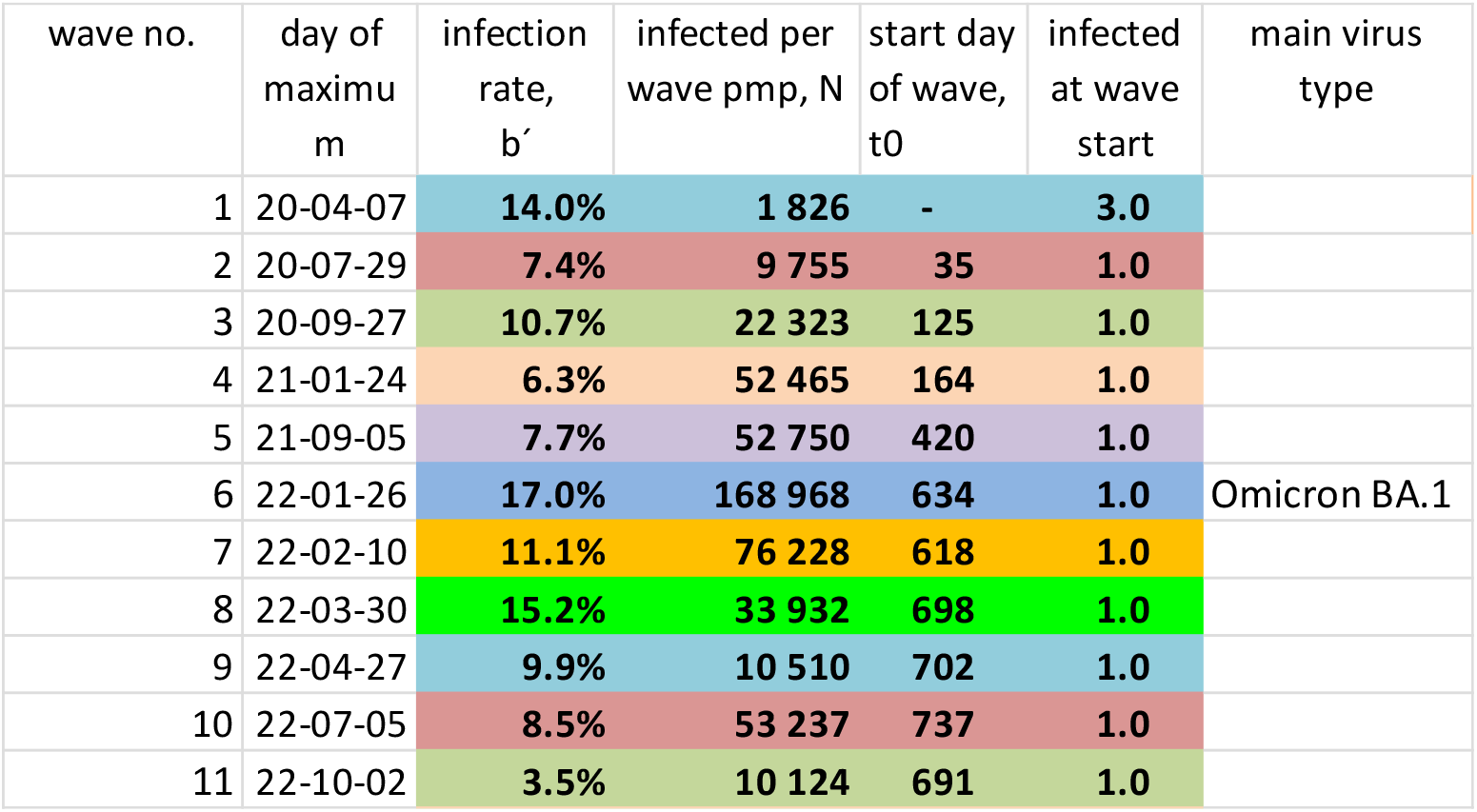
Israel, SI parameters

**Fig. A10.**
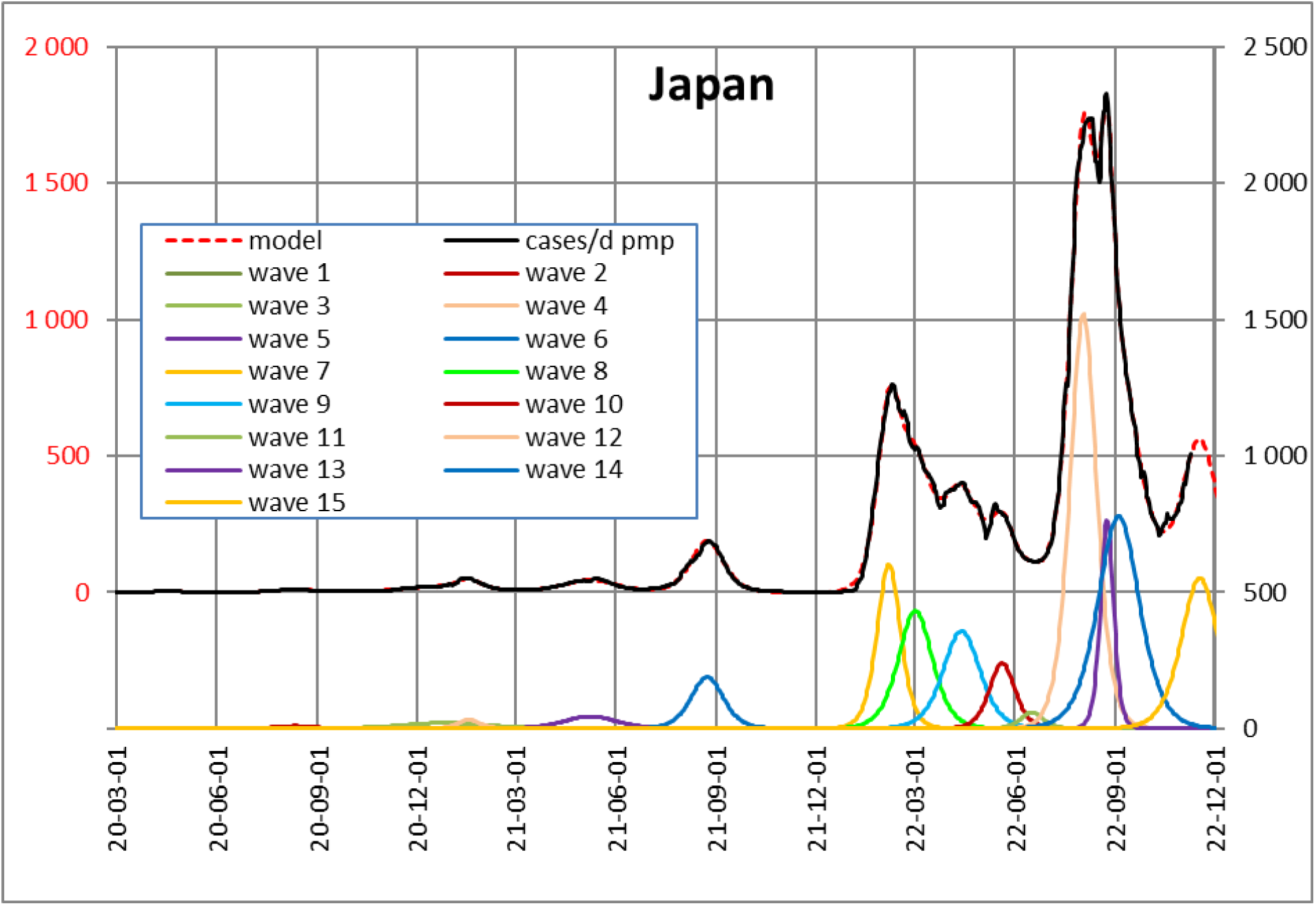
Japan, real course and models

**Table A10.** Japan, SI parameters

| wave no. | day of maximum | infection rate, $b'$ | infected per wave pmp, N | start day of wave, $t_0$ | infected at wave start | main virus type |
| --- | --- | --- | --- | --- | --- | --- |
| 1 | 20-04-18 | 12.1% | 132 |  | 0.1 |  |
| 2 | 20-08-12 | 8.4% | 439 | 100 | 1.0 |  |
| 3 | 20-12-26 | 3.9% | 2 127 | 112 | 1.0 |  |
| 4 | 21-01-17 | 14.9% | 875 | 286 | 1.0 |  |
| 5 | 21-05-08 | 6.2% | 2 875 | 313 | 1.0 |  |
| 6 | 21-08-23 | 10.1% | 7 486 | 460 | 1.0 |  |
| 7 | 22-02-06 | 13.9% | 17 257 | 645 | 1.0 | Omicron BA.1 |
| 8 | 22-03-02 | 9.6% | 17 963 | 638 | 1.0 |  |
| 9 | 22-04-13 | 8.9% | 16 104 | 673 | 1.0 |  |
| 10 | 22-05-20 | 12.8% | 7 523 | 749 | 1.0 |  |
| 11 | 22-06-17 | 14.4% | 1 633 | 795 | 1.0 |  |
| 12 | 22-08-03 | 12.4% | 49 048 | 806 | 1.0 |  |
| 13 | 22-08-24 | 24.9% | 12 257 | 877 | 1.0 |  |
| 14 | 22-09-04 | 7.9% | 39 620 | 791 | 1.0 |  |
| 15 | 22-11-17 | 8.8% | 25 013 | 885 | 1.0 |  |

**Fig. A11.**
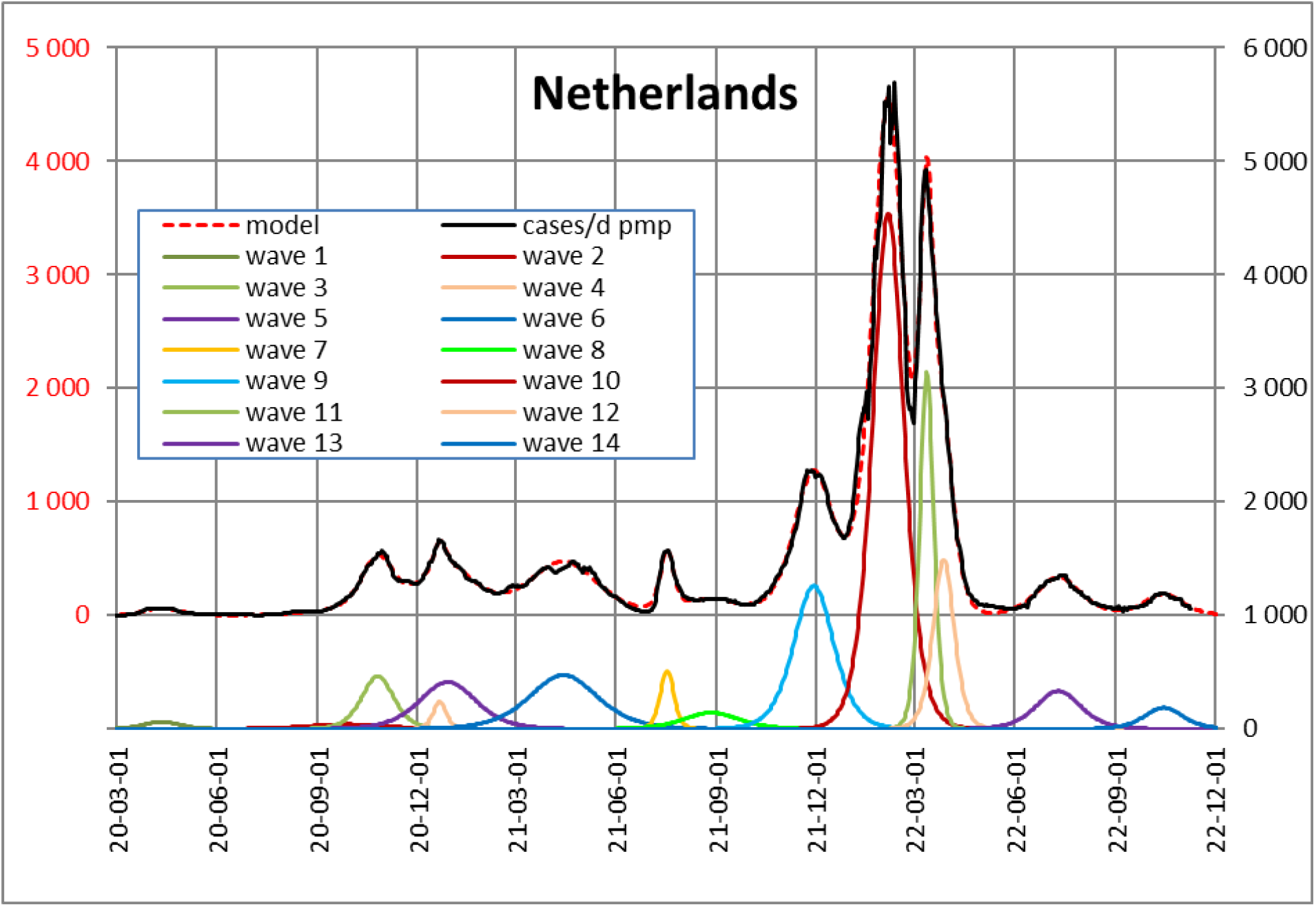
Netherlands, real course and models

**Table A11.**
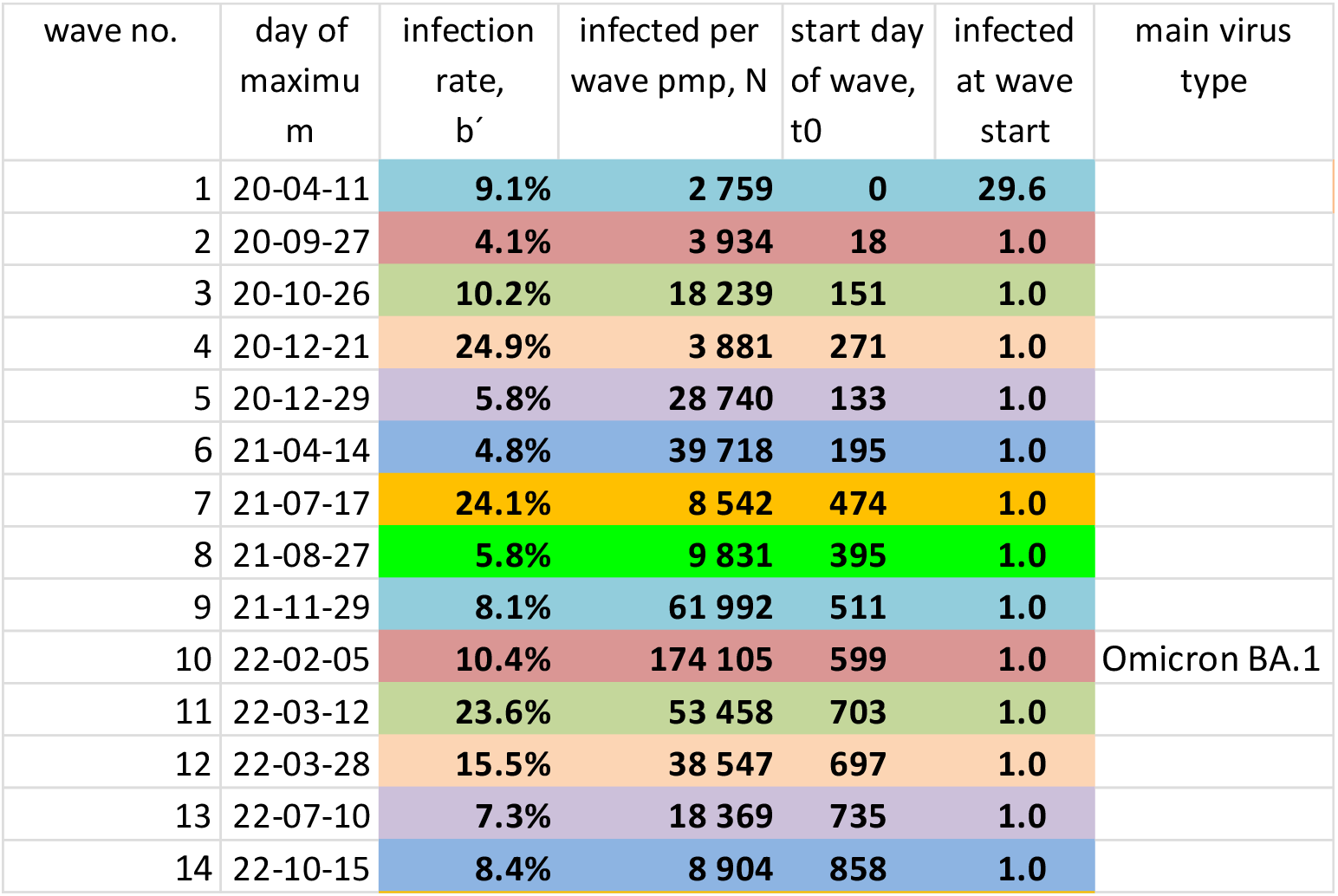
Netherlands, SI parameters

| wave no. | day of maximum | infection rate, $b'$ | infected per wave pmp, N | start day of wave, $t_0$ | infected at wave start | main virus type |
| --- | --- | --- | --- | --- | --- | --- |
| 1 | 20-04-11 | 9.1% | 2 759 | 0 | 29.6 |  |
| 2 | 20-09-27 | 4.1% | 3 934 | 18 | 1.0 |  |
| 3 | 20-10-26 | 10.2% | 18 239 | 151 | 1.0 |  |
| 4 | 20-12-21 | 24.9% | 3 881 | 271 | 1.0 |  |
| 5 | 20-12-29 | 5.8% | 28 740 | 133 | 1.0 |  |
| 6 | 21-04-14 | 4.8% | 39 718 | 195 | 1.0 |  |
| 7 | 21-07-17 | 24.1% | 8 542 | 474 | 1.0 |  |
| 8 | 21-08-27 | 5.8% | 9 831 | 395 | 1.0 |  |
| 9 | 21-11-29 | 8.1% | 61 992 | 511 | 1.0 |  |
| 10 | 22-02-05 | 10.4% | 174 105 | 599 | 1.0 | Omicron BA.1 |
| 11 | 22-03-12 | 23.6% | 53 458 | 703 | 1.0 |  |
| 12 | 22-03-28 | 15.5% | 38 547 | 697 | 1.0 |  |
| 13 | 22-07-10 | 7.3% | 18 369 | 735 | 1.0 |  |
| 14 | 22-10-15 | 8.4% | 8 904 | 858 | 1.0 |  |

**Fig. A12.**
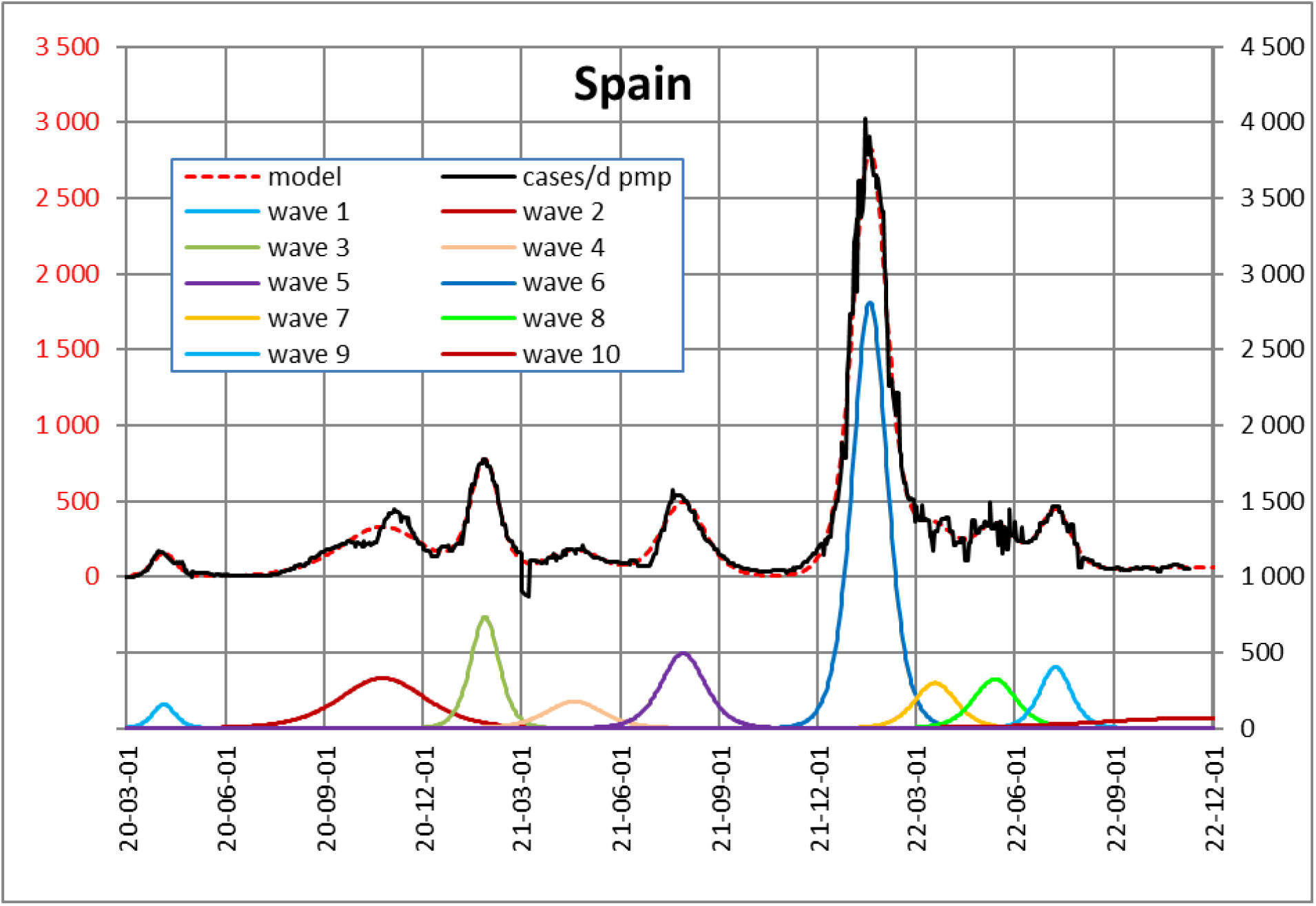
Spain, real course and models

**Table A12.** Spain, SI parameters

| wave no. | day of maximum | infection rate, $b'$ | infected per wave pmp, N | start day of wave, $t_0$ | infected at wave start | main virus type |
| --- | --- | --- | --- | --- | --- | --- |
| 1 | 20-04-05 | 14.4% | 4 395 |  | 8.7 |  |
| 2 | 20-10-24 | 3.8% | 35 154 |  | 3.5 |  |
| 3 | 21-01-26 | 11.3% | 25 884 | 250 | 1.0 |  |
| 4 | 21-04-19 | 5.7% | 12 347 | 258 | 1.0 |  |
| 5 | 21-07-29 | 7.3% | 26 858 | 384 | 1.0 |  |
| 6 | 22-01-17 | 9.3% | 121 113 | 570 | 1.0 | Omicron BA.1 |
| 7 | 22-03-19 | 7.7% | 15 459 | 632 | 1.0 |  |
| 8 | 22-05-13 | 7.6% | 17 054 | 683 | 1.0 |  |
| 9 | 22-07-08 | 10.3% | 15 775 | 773 | 1.0 |  |
| 10 | 22-11-08 | 1.7% | 15 007 | 425 | 1.0 |  |

**Fig. A13.**
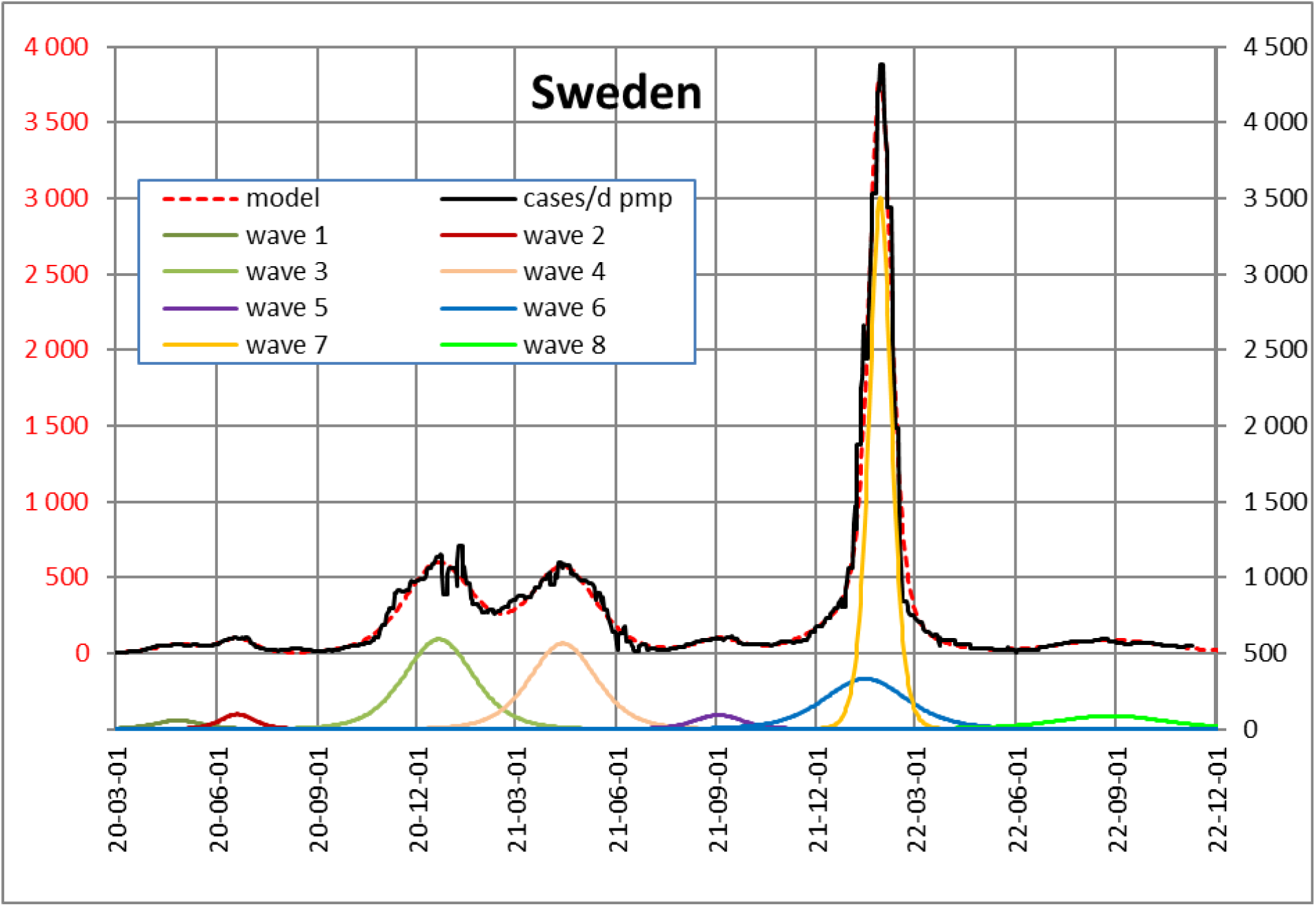
Sweden, real course and models

**Table A13.** Sweden, SI parameters

|  |  |  |  |  |  |  |
| --- | --- | --- | --- | --- | --- | --- |
| 1 | 20-04-26 | 7,0% | 3.210 | - | 33,7 |  |
| 2 | 20-06-20 | 9,6% | 4.015 | - | 0,0 |  |
| 3 | 20-12-21 | 4,7% | 49.953 | 75 | 1,0 |  |
| 4 | 21-04-13 | 4,9% | 45.740 | 199 | 1,0 |  |
| 5 | 21-09-02 | 6,8% | 5.347 | 433 | 1,0 |  |
| 6 | 22-01-15 | 4,0% | 33.360 | 431 | 1,0 |  |
| 7 | 22-01-28 | 14,8% | 94.485 | 629 | 1,0 | Omicron BA.1 |
| 8 | 22-08-27 | 3,0% | 11.633 | 602 | 1,0 |  |

**Fig. A14.**
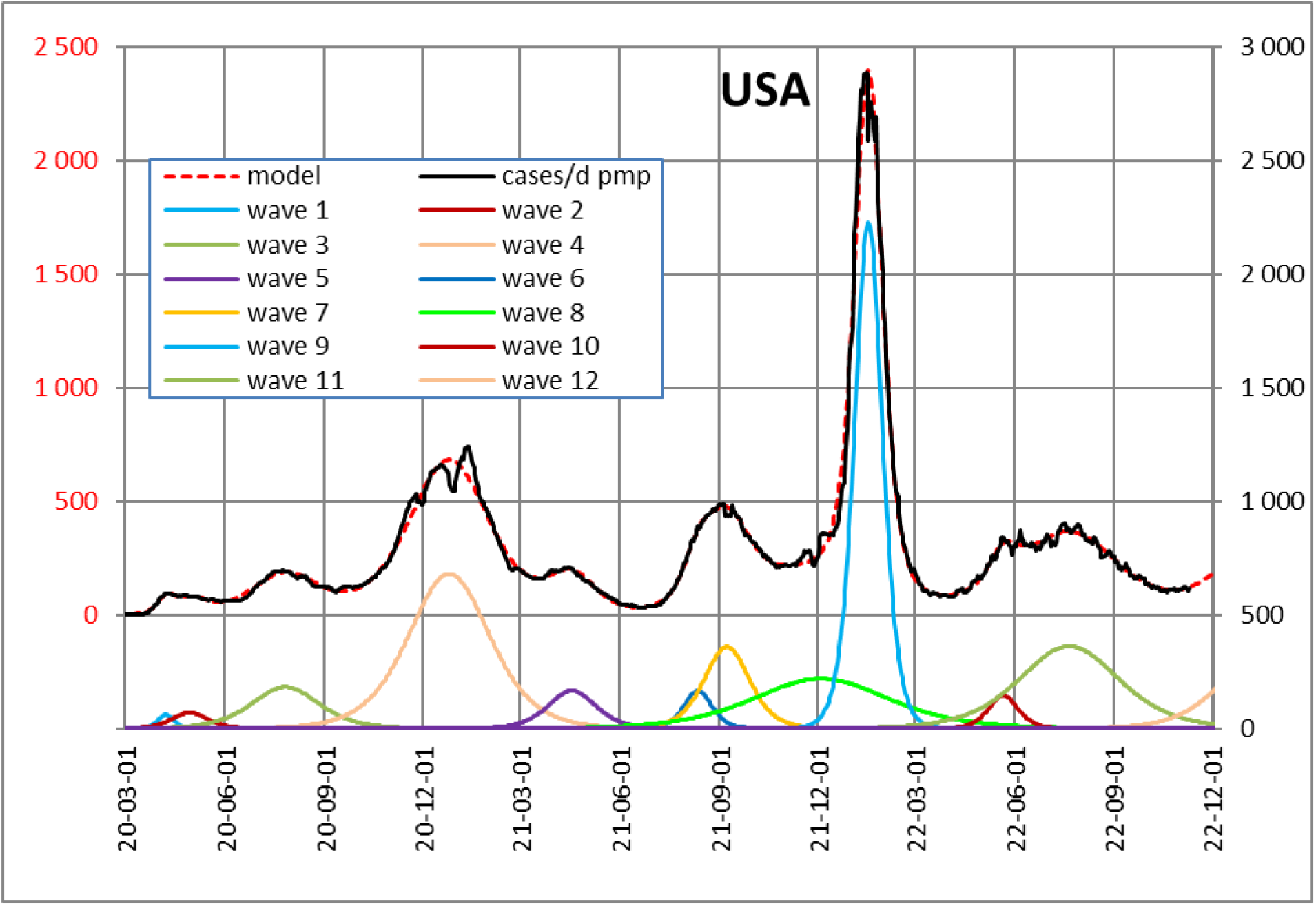
USA, real course and models

**Table A14.** USA, SI parameters

| wave no. | day of maximum | infection rate, $b'$ | infected per wave pmp, N | start day of wave, $t_0$ | infected at wave start | main virus type |
| --- | --- | --- | --- | --- | --- | --- |
| 1 | 20-04-07 | 20.4% | 1 201 | - | 0.1 |  |
| 2 | 20-04-29 | 9.1% | 3 099 | - | 6.6 |  |
| 3 | 20-07-27 | 4.8% | 15 328 | - | 9.1 |  |
| 4 | 20-12-25 | 4.0% | 68 675 | 27 | 1.0 |  |
| 5 | 21-04-18 | 6.8% | 9 884 | 285 | 1.0 |  |
| 6 | 21-08-12 | 11.4% | 5 788 | 462 | 1.0 |  |
| 7 | 21-09-08 | 7.1% | 20 245 | 425 | 1.0 |  |
| 8 | 21-12-04 | 2.4% | 36 797 | 210 | 1.0 |  |
| 9 | 22-01-17 | 11.7% | 76 324 | 599 | 1.0 | Omicron BA.1 |
| 10 | 22-05-21 | 10.8% | 5 384 | 741 | 1.0 |  |
| 11 | 22-07-21 | 3.2% | 44 862 | 549 | 1.0 |  |
| 12 | 23-01-09 | 4.0% | 28 627 | 798 | 1.0 |  |

**Fig. A15.**
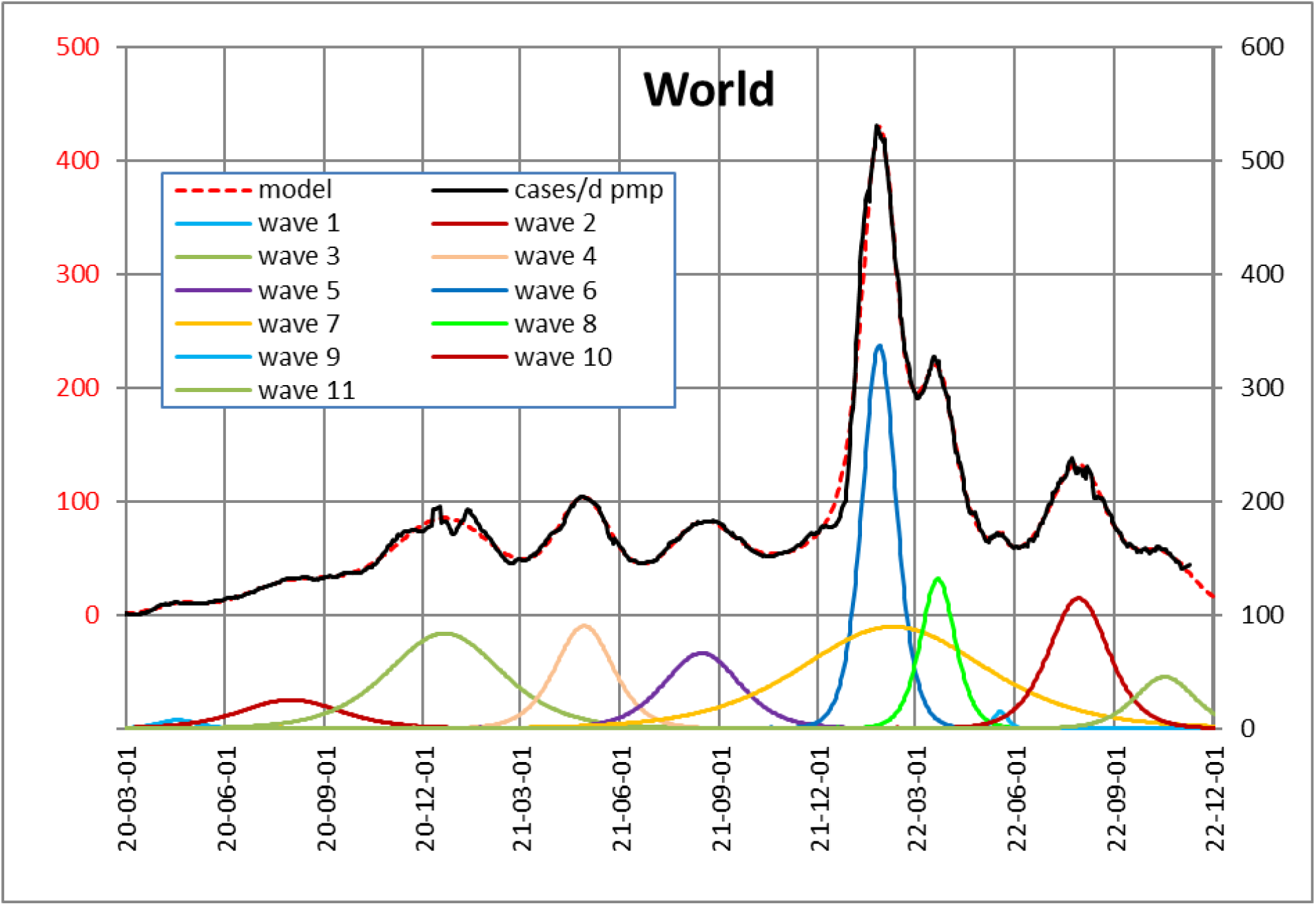
World, real course and models

**Table A15.** World, SI parameters

| wave no. | day of maximum | infection rate, $b'$ | infected per wave pmp, N | start day of wave, $t_0$ | infected at wave start | main virus type |
| --- | --- | --- | --- | --- | --- | --- |
| 1 | 20-04-17 | 8.2% | 359 | - | 3.6 |  |
| 2 | 20-07-31 | 3.2% | 3 139 | - | 18.0 |  |
| 3 | 20-12-21 | 2.8% | 11 813 | - | 2.2 |  |
| 4 | 21-04-28 | 5.6% | 6 470 | 275 | 1.0 |  |
| 5 | 21-08-15 | 4.3% | 6 222 | 336 | 1.0 |  |
| 6 | 22-01-26 | 9.6% | 14 111 | 605 | 1.0 | Omicron BA.1 |
| 7 | 22-02-08 | 1.8% | 20 455 | 151 | 1.0 |  |
| 8 | 22-03-21 | 9.3% | 5 651 | 666 | 1.0 |  |
| 9 | 22-05-18 | 23.4% | 254 | 793 | 1.0 |  |
| 10 | 22-07-30 | 5.4% | 8 476 | 722 | 1.0 |  |
| 11 | 22-10-17 | 5.4% | 3 371 | 818 | 1.0 |  |

## Notes

### Competing Interest Statement

The authors have declared no competing interest.

### Funding Statement

This study did not receive any funding.

